# Domain specific phenotypic expansion associated with variants in *MACF1*

**DOI:** 10.1101/2025.06.26.25330137

**Authors:** Nikhita Gogate, Angad Jolly, Jill A. Rosenfeld, Paulina Bahena-Carbajal, Jonathan A. Bernstein, Devon Bonner, Tiffany Busa, Ingrid Cristian, Precilla D’Souza, Jennifer Friedman, Svetlana Gorokhova, Thomas Haaf, Isabella Herman, Ugur Ufuk Isin, Shalini N Jhangiani, Ivy Johnson, Jerica Lenberg, Ellen F. Macnamara, Reza Maroofian, Undiagnosed Diseases Network, Melissa Racobaldo, Olivia L Redlich, Cynthia Tifft, Tulay Tos, Barbara Vona, Regina M. Zambrano, Ingrid M. Wentzensen, Kristen Wigby, Davut Pehlivan, Richard A Gibbs, James R Lupski, Jennifer E Posey

**Author notes:** Correspondence: Jennifer Ellen Posey, M.D., Ph.D. Columbia University Irving Medical Center 701 W 168th St, 14-1406, New York, NY 10032.

## Abstract

**Purpose:** While heterozygous *de novo* missense variants in the microtubule-binding GAR domain of Microtubule-actin cross-linking factor 1 (*MACF1*) cause Lissencephaly 9 with Complex Brainstem Malformations [MIM #618325], the phenotypic impact of variants outside this domain remains unclear.

**Methods:** Through collaborative efforts, we assembled a cohort of 10 affected individuals from 8 unrelated families with either biallelic or monoallelic non-GAR domain *MACF1* variants who exhibit partially overlapping yet unique phenotypic traits. Combined with previously reported cases, we analyzed genotype and phenotype data from 29 individuals using Human Phenotype Ontology (HPO)-based unsupervised hierarchical clustering.

**Results:** Clustering revealed two distinct phenotypic signatures, suggesting domain-specific effects. Variants outside the GAR domain associate with broader neurodevelopmental phenotypes and variable craniofacial and skeletal expressivity. Additionally, enrichment analysis (p < 0.001) using OMIM HPO sets supported these findings. In contrast to the GAR domain’s strong correlation with lissencephaly and brainstem malformations, biallelic non-GAR domain *MACF1* variants were linked to diverse developmental anomalies.

**Conclusion:** These results expand the phenotypic spectrum of *MACF1*-related disorders and highlight the relevance of domain-specific variant effects. Comprehensive genetic and phenotypic assessments are essential for understanding the role of *MACF1* in development, informing diagnosis, and guiding future research on cytoskeletal regulation in neurodevelopment.

## Introduction

*MACF1* encodes <u>M</u>icrotubule-<u>A</u>ctin <u>C</u>ross-linking <u>F</u>actor 1, a highly versatile protein known for its crucial role in cytoskeletal organization and dynamics. In humans, *MACF1* is located on chromosome 1p34.3, spanning ∼300kb and comprising 101 exons^1,2^. The full-length protein spans ∼7555 amino acids, with an approximate molecular weight of 860 kDa, making it one of the larger cytoskeletal proteins. A modular domain structure that supports its critical roles in cellular organization, migration, and signaling^3^. At the N-terminus, there are two calponin homology (CH) domains constitute the functional actin-binding domain, enabling direct interaction with actin filaments, and anchoring the protein to the actin cytoskeleton and facilitating its integration with other cellular components^4,5^. The central portion of MACF1 contains the plakin domain and spectrin repeats, defining MACF1 as a member of the spectraplakin protein family. The plakin domain mediates interactions with cellular junctions and the cortical cytoskeleton, while the spectrin repeats confer structural flexibility and serve as scaffolding elements for protein-protein interactions necessary to maintain cytoskeletal integrity^1,2,4,6^. EF-hand calcium binding motifs downstream of the spectrin repeats provide regulatory functions by modulating the activity of MACF1 in response to calcium signaling. The C-terminus of MACF1 harbors a growth arrest specific 2 (GAS/GAR) domain that allows MACF1 to directly bind to microtubules, contributing to crosslinking activities^2,4^. This intricate domain arrangement allows MACF1 to act as a “mega-scaffold,” linking cytoskeletal elements and coordinating their functions with intracellular signaling pathways. MACF1 functions are pleiotropic, having been implicated as a regulator of the GSK3β/TCF7^7,8^ control of migration during bone development, a regulator of skeletal muscle function through cytoskeletal-based localization of myonuclei^9^, and perhaps most notably as a critical gene in cortical development through its role in the cell polarity and migration of neural progenitors and axon guidance^7–12^.

The multifunctional role of MACF1 in cellular architecture and signaling explains its association with a diverse array of developmental phenotypes. A 2014 study described a family with neuromuscular features including hypotonia, reduced motor coordination, joint contractures, and low muscle tone. Affected individuals harbored a 302kb heterozygous duplication on 1p34.3 encompassing exons 1 through 36 and part of exon 37 at the N-terminus of *MACF1*. Functional studies showed decreased *MACF1* mRNA and protein expression^13^. In 2018, Dobyns et al. reported nine probands with recurrent *de novo* heterozygous missense variants affecting the zinc ion-binding pocket in the GAR domain of MACF1^14^. These variants were shown to impair neuronal migration and axon guidance through dominant-negative mechanisms, resulting in a distinct phenotype characterized by lissencephaly and a W-shaped brainstem malformation caused by hypo– or aplasia of the pontine crossing fibers^14^. Bölsterli et al. (2021) further proposed that this MRI pattern is pathognomonic for *MACF1* variants in the GAR domain^15^. However, several additional studies have expanded the phenotypic spectrum associated with *MACF1* variants outside the GAR domain to variably include infantile spams and epilepsy^16^, hypotonia and muscle weakness^17^, congenital myasthenic syndromes with limb-girdle weakness^11,18^, non-syndromic hearing loss^19^, autism spectrum disorder^20^, structural brain malformations^21^. Rare *MACF1* variants have also been linked to neurobehavioral and neurodegenerative disorders such as schizophrenia, bipolar disorder^22–24^ and Parkinson’s disease, where its role in cytoskeletal stability and cellular transport pathways may contribute to pathogenesis^25^. These findings emphasize the multifunctional role of *MACF1* in maintaining cellular and tissue integrity across systems, with variants disrupting its scaffolding and signaling capabilities. As genomics efforts advance, further studies will likely uncover additional roles for *MACF1* in rare and common diseases, broadening our understanding of its contribution to human pathology.

In this study, a collaborative effort facilitated by matchmaking of geneticists interested in genotype-phenotype studies (GeneMatcher^26^ and Undiagnosed Diseases Network^27^) allowed for the independent identification of 10 affected individuals from 8 families with segregating variants mapping closer to the N-terminus of *MACF1*. These variant alleles affected or were adjacent to actin binding and spectrin domains rather than the microtubule binding GAR domain, and affected individuals had a phenotype that included microtia, facial asymmetry, microcephaly, and spasticity. To better delineate the domain-specific phenotypic spectrum of MACF1, quantitative phenotypic analyses using the Human Phenotype Ontology (HPO)^28^ were performed. Our data suggest that both mono– and biallelic variants outside the GAR-domain of MACF1 may indeed cause a partially overlapping but unique disease trait distinct from that observed in individuals with monoallelic variants within the GAR domain. Furthermore, the observed phenotype-genotype association implicates *MACF1* in yet another potential role in neurodevelopment and craniofacial phenotypes.

## Methods

### Identification of individuals with *MACF1* variants

The study was conducted with approval from the Baylor College of Medicine (BCM) Institutional Review Board. Written, informed consent was obtained from all participants or their guardians under BCM protocol H-29697 or through other institutional review boards at each respective institution. Participants were identified via the Baylor-Hopkins Center for Mendelian Genomics (BHCMG) / BCM Genomics Research Elucidates the Genetics of Rare disease (BCM-GREGoR) database, the Baylor Genetics (BG) clinical diagnostic laboratory database, the Undiagnosed Diseases Network (UDN)^27^, GeneMatcher^26^, or other research and clinical diagnostic laboratories. All participants were evaluated by a clinical geneticist and/or neurologist. Detailed pedigrees and phenotypic data were collected from collaborating clinicians using a standardized template.

### Exome/Genome sequencing and analysis

Sequencing methods are detailed in the Supplemental Text. We considered all *MACF1* variants identified in our cohort for inclusion based on phenotypic consistency, regardless of their clinical classification. All rare, predicted damaging single-nucleotide variants and indels were reviewed in accordance with ACMG/AMP guidelines to ensure standardized variant interpretation^29^ (Supplemental table 1).

### Human Phenotype Ontology (HPO)-based quantitative phenotypic similarity analysis

After ascertainment of a cohort of individuals with putatively damaging variation of *MACF1*, the clinical data obtained from all cases were annotated with Human Phenotype Ontology (HPO) terms using RAG-HPO^30^ followed by manual curation. The number of HPO terms assigned to each proband was used to assess the depth of phenotypic annotation within the cohort (Figure S1), with an average of 16.7 HPO terms/case. To ensure sufficient resolution for quantitative phenotypic analysis, a minimum of four HPO terms per case was required for inclusion. This threshold was selected based on the premise that comprehensive phenotypic annotation improves the reliability of semantic similarity computations and facilitates the identification of distinct and biologically meaningful clusters within the dataset. To perform the quantitative phenotype comparison analyses, HPO term sets from each newly ascertained individual and each individual previously reported to have a GAR domain variant were compared in a pairwise fashion. These case term sets were also compared to the publicly available HPO annotated OMIM gene (4869) and disease (8384) associated phenotype database (https://hpo.jax.org/data/annotations).

To perform ontological similarity comparison, a symmetric Lin score was generated as provided in the OntologyX^31^ suite of R packages. P-values were calculated by comparing the Lin similarity^32^ of a proband phenotype and a matching gene or disease to that of 100,000 randomly selected pairs from all term sets being compared. A p-value cutoff of 0.001 was used to define the set of most similar gene associated phenotype sets or disease phenotype sets to include in further analyses. The group of MACF1 probands were clustered with matching genes or disease phenotype sets. A gap statistical analysis, where goodness of clustering was measured against number of clusters assigned, allowed definition of an optimal number of clusters (Figure S2). Hierarchical Agglomerative Clustering using the Ward method was then applied to cluster the clinically observed phenotypes of the patients, and results visualized on a heatmap of pairwise Lin phenotypic similarity score using the R package ComplexHeatmap.

## Results

### Identification of cases

The index case (Individual 1, Family 1) presents with facial asymmetry, facial hyperostosis and left central facial paralysis. Notable features include small crumpled and cupped ears, microstomia and small oral aperture, 5^th^ finger clinodactyly, hirsutism and synophrys (Figure 1, Table 1). Brain MRI showed right frontotemporal cortical dysplasia, along with calcification of the falx cerebri. Sleep-wake video EEG showed focal epileptic findings in the left frontocentral region. She was born to consanguineous parents. Trio-based ES prioritized a homozygous *MACF1* variant [NM_001394062.1:c.4069G>A, p.(E1357K)] within an Absence of Heterozygosity (AOH) region of 19.9Mb (Total AOH 146.8Mb) (Figure S3). To comprehensively define the phenotypic spectrum of *MACF1* with possible domain specificity, we leveraged the online matchmaking platform GeneMatcher^26^ and explored additional datasets from research and diagnostic laboratories such as the UDN^27^ and Baylor Genetics diagnostic laboratory. We identified 10 affected individuals from 8 unrelated families with neurodevelopmental disorders who carry rare biallelic or monoallelic variants in *MACF1* located outside the GAR domain (Figure 1, Table 1). These variants are predicted to be damaging by in silico tools (CADD PHRED scores >20^33^) and are located in highly conserved residues (GERP++ >5^34^). Additional rationale for inclusion of specific cases and variants is provided in the Supplemental Text.

**Figure 1:**
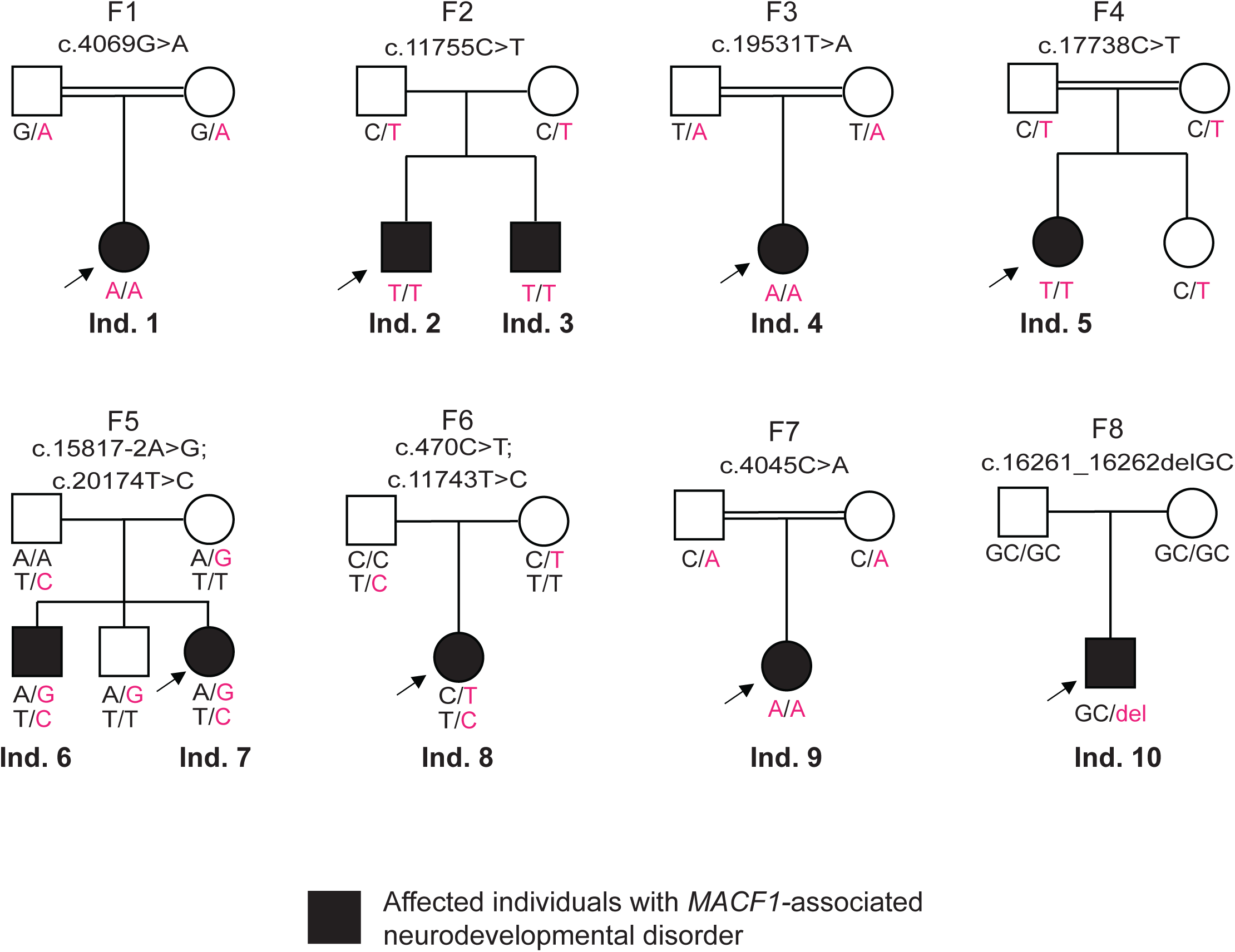
Pedigrees and their genotypes for families with *MACF1* variants. Standard pedigree structures are utilized—filled circles and squares denote clinically affected individuals, and probands are indicated by black arrows. Nine families with 11 individuals affected with *MACF1*-related neurodevelopmental disorders that are identified through the BHCMG/ BCM-GREGoR, the Baylor Genetics (BG) clinical diagnostic laboratory database, Undiagnosed Diseases Network (UDN), and GeneMatcher. All participants were evaluated by a clinical geneticist and/or neurologist. Detailed pedigrees and phenotypic data were collected from collaborating clinicians using a standardized template.

**Table 1:** Summary of clinical phenotypic data for individuals identified in this study.

|  | Family 1 |
| --- | --- |
|  | Individual 1 |
| Nucleotide (NM_001394062.1) | c.4069G>A |
| Protein | p.E1357K |
| Zygoty | Hom |
| Consanguinity | Yes |
| Sex | F |
| Head circumference cm(%) (SD) | 35 percentile (-0.38 SD) |
| Head and neck anomalies | Hirsutism, Synophrys |
| Hemifacial Microsomia/Facial Asymmetry (Uni or bilateral) | + |
| Hemifacial atrophy | - |
| Facial palsy/paralysis | + |
| Micrognathia | - |
| Preauricular tags | + |
| Microtia | + |
| Hearing loss | + |
| Microcephaly | - |
| Cleft palate | - |
| Dental anomalies | - |
| Skeletal abnormalities | Clinodactyly 5th toes |
| Neurologic symptoms | - |
| Seizures | - |
| Global developmental delay | - |
| Speech delay | - |
| Motor delay | - |
| Intellectual disability | - |
| MRI Craniofacial/Neurologic Finding | Cortical dysplasia (thick cortex, R frontotemporal), calcification of falx cerebri |
| Other findings | Focal epileptic findings (Individual has no seizure) |

| Family 2 |  | Family 3 |
| --- | --- | --- |
| Individual 2 | Individual 3 | Individual 4 |
| c.11755C>T | c.11755C>T | c.19531T>A |
| p.R3919W | p.R3919W | p.S6511T |
| Hom | Hom | Hom |
| - | - | Yes |
| M | M | F |
|  |  | < 1 percentile (-3.10 SD) |
| Small forehead, Smaller right eye, High palate, Tubular nose, Long smooth philtrum, Asymmetric crying facies noted | High palate, Long smooth philtrum | Narrow face, prominent and cupped ears, simple and large nose, full lips, synophrys, hirsutism |
| + | - | - |
| + | - | - |
| + (unilateral) | + (unilateral) | - |
| + | + | + |
| - | - | - |
| + | + | - |
| + | + | + (conductive) |
| + | + | + |
| - | - | + |
| - | - | - |
| Clinodactyly, Small 4th/5th toes | - | Bilateral forearms demonstrate the left radial head to be posteriorly dislocated congenitally with a shallow capitellum. The left ulna is positive in relationship to the radius at the wrist level. Left arm does not supinate. Short 5th metatarsals, short distal phalanges. |
| - | - | Progressive functional and cognitive decline (now non-verbal, non-ambulatory, requiring g-tube, trach)<br>Hyperactivity, generalized hypotonia, generalized weakness, spasticity, episodic dyskinesias, Abnormal EEG not associated with abnormal movements bells phenomenon, Lagophthalmos |
| - | - | - |
| - | - | + (severe) |
| - | - | + |
| - | - | + |
| - | - | + |
| Thin corpus callosum, cerebral white matter atrophy | Mild periventricular foci of increased FLAIR and T2 signal | Progressive global brain atrophy (includes cerebrum, deep gray nuclei, cerebellum, brainstem, corpus callosum) particularly notable ,in the pons and cerbellum; hot-cross bun sign |
| Spastic diplegia<br>Periventricular leukomalacia |  | Pre and post natal growth deficiency with short stature.<br>Bulbous toes. On hands, single palmar crease bilaterally, decreased thenar crease on right with absent thenar crease on left with striking thenar hypoplasia, no creases on left thumb.<br>Early puberty;<br>Hypoplastic toe nails 4-5;<br>Cataracts.<br>Muscle biopsy showed mild myopathic changes.<br>In addition, large deletion on muscle mtDNA m.3259-16069 at 40% heteroplasmy which is absent in blood and buccal mtDNA, and homozygous variant in gene of uncertain significance PARQ3 (c.864G>C; pW288C) |

| Family 4 | Family 5 |  |
| --- | --- | --- |
| Individual 5 | Individual 6 | Individual 7 |
| c.17738C>T | c.15817-2A>G; c.20174T>C | c.15817-2A>G; c.20174T>C |
| p.P5913L | p.L6725P | p.L6725P |
| Hom | Comp. het | Comp. het |
| No | No | No |
| F | M | F |
| < 1 percentile (-2.99 SD) | 36 percentile (-0.36 SD) | 17 percentile (-0.95 SD) |
| Prominent forehead, small head | - | - |
| - | - | - |
| - | - | - |
| - | - | - |
| - | - | - |
| - | - | - |
| - | - | - |
| - | - | - |
| + | - | - |
| - | - | - |
| Melanosis of the gums | - | - |
| clinodactyly of fingers 3,4 and 5 and the 2nd and 4th toes overlap the third. The second and fourth metacarpals in both hands are shortened as compared to the first third and fifth. The third and fourth metatarsals in both feet are shortened as compared to the first second and fifth. | - | - |
|  | Autism spectrum disorder;<br>paroxysmal abnormal movements | Gross motor delay, hypotonia, tethered spinal cord |
| - | Normal EEG in context of abnormal movements | - |
| + | - | - |
| + | - | - |
| + | - | + |
| - | Not evaluated | Not evaluated |
| Had MRI, No intracranial findings. the dens been somewhat posteriorly, and there is partial fusion and hypoplasia of C6 and C7. So-called posterior vertebral scalloping (concave posterior vertebral body) is noted in L1, L2, L3, and L4 vertebral bodies. | MRI brain with nonspecific T2/FLAIR hyperintensity. |  |
| Mild OSA, had T&A |  |  |

| Family 6 | Family 7 | Family 8 |
| --- | --- | --- |
| Individual 8 | Individual 9 | Individual 10 |
| c.470C>T, c.11743T>C | c.4045C>A | c.16261_16262del |
| p.T157I, p.S3915P | p.L1349M | p.Q5421Afs*5 |
| Comp. het | Hom | Het |
|  | Yes | No |
| F | F | M |
| < 1 percentile (-4.62 SD) | 1 percentile (-2.29 SD) | < 1 percentile (-3.11 SD) |
| Slanting forehead, flat nasal bridge, smooth philtrum, short webbed neck, frontal parietal plagiocephaly, left facial drooping, synophrys and retrognathia | Cleft palate, large tongue | - |
| + (while smiling) | + | - |
| - | + | - |
| - | - | - |
| + | + | - |
| - | - | - |
| + | - | - |
| + (conductive) | + (bilateral) | - |
| + | + | + |
| + | + | - |
| Primary delayed dentition | - | - |
| brachydactyly, syndactyly of second and third toe, Skeletal dysplasia<br>Short foot<br>Short toe<br>Humeroradial synostosis | Club foot | - |
| Generalized hypotonia<br>Spasticity<br>EEG abnormality | + | + |
|  | - | + |
| + | + | + |
| + | + | + |
| + | + | + |
| - | + | + |
| No parenchymal abnormality identified. Mild prominence of the extra-axial CSF spaces | No | Left cortical frontal dysplasia |
| feeding difficulties, poor weight gain, gastrostomy tube feed dependence, failure to thrive, Bilateral ptosis, Speech apraxia, Ankyloglossia | poor feeding, dysphagia, recurrent aspirations, bilateral club foot were identified and assisted. Posterior clinical examinations revealed short stature, poor weight, severe developmental delay without regression, generalized muscle hypotonia and ataxia. Auricular septal defect | coarse features, type 1 diabetes, behaviour trouble with treatment, able to read a few words, EEG : abnormal epileptiform activity but of moderate abundance in the left frontotemporal region. |

We analyzed genotype and phenotype data from these 10 cases, as well as previously reported disease-associated *MACF1* variants included in this study (Table S2)^11,14,15,17,18,20,21,35^. These cases display a wide phenotypic spectrum, spanning severe structural brain anomalies such as lissencephaly and cortical dysplasia, as well as neuromuscular features and diverse neurodevelopmental and neurological manifestations. Collectively, these cases inform a systematic assessment of the role of both biallelic and monoallelic *MACF1* variants outside the GAR domain in contributing to neurodevelopmental phenotypes.

### Domain specificity and *MACF1* variants

The molecular and genetic data regarding the variants and families are summarized in Table 2. More detailed clinical and molecular data are described in Table S1. Throughout this report, all sequence variants described are based on MANE select reference sequence GenBank: NM_001394062.1. In our cohort, a total of five families with homozygous variants, two families with compound heterozygous variants, and one family with heterozygous variant were identified. These include eight unique missense variants, one splice-site variant and one frameshift variant, which are collectively categorized among the above classifications described. MACF1 has 5 different domains that interact with various molecules and support its critical roles in cellular organization, migration, and signaling^4^. Mapping the variants identified in our cohort, we observe domain-specific phenotypes (Figure 2). Variants in the GAR domain are associated with characteristic structural brain abnormalities, including a W-shaped brainstem, dysplastic cerebellum, and thin corpus callosum^14^. In contrast, variants in other MACF1 domains result in a partially overlapping yet broader phenotypic spectrum, encompassing craniofacial and skeletal anomalies such as microcephaly, cleft palate, micrognathia, and clinodactyly. The different phenotypes associated with these regions reflect the multiple roles and domain functions of MACF1 in cytoskeletal structure and cellular signaling.

**Figure 2:**
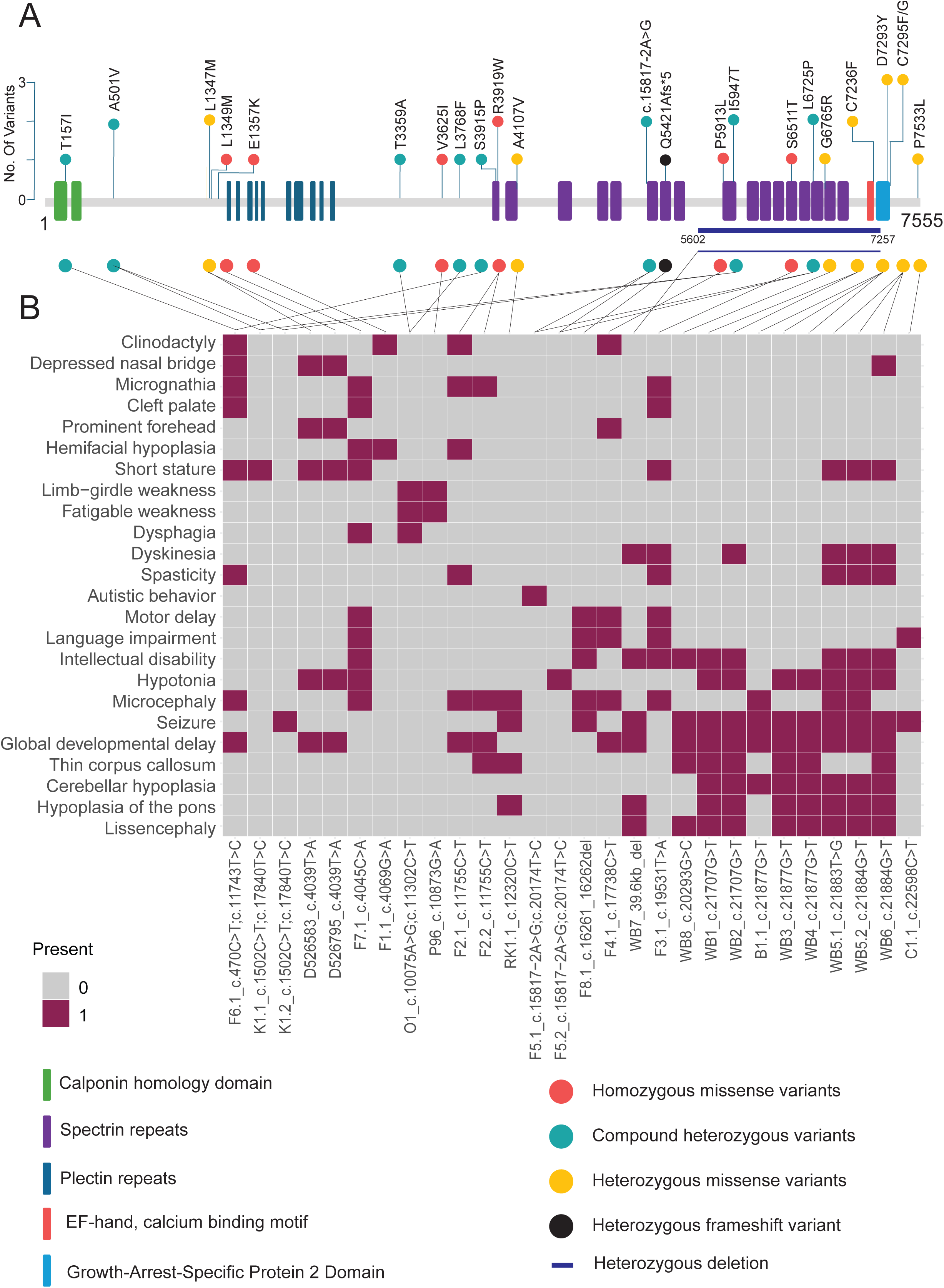
Schematic diagram of MACF1 protein structure. (A) Conceptual representation of MACF1 protein structure produced by translation of MANE transcript NM_001394062.1 with mapping location (circles) of the amino acid variants observed in this study. In total, we identified 23 different SNVs and indels that are distributed throughout the protein. A larger deletion (39.6kb) is shown in a dark blue line. Homozygous missense variants are represented in orange circles, while heterozygous missense variants are denoted in yellow. Greenish blue circles represent compound heterozygous variants, and the black circle is for heterozygous frameshift variant observed in this study. (B) Annotation grid of phenotypes highlighting domain specificity. Columns represent individual cases, mapped to the MACF1 protein based on the identified variant. Phenotype terms are listed in each row. Variants within the GAR domain are associated with Lissencephaly and cerebellar hypoplasia, whereas variants outside the GAR domain correspond to a broader neurodevelopmental phenotype, including craniofacial and skeletal abnormalities.

**Table 2:** Genotype summary of individuals identified in this study.

| Individual | Zygoty | Genomic Location (hg38) | Nucleotide/Protein (NM_001394062.1) | Minor Allele frequency / Hmz count (gnomAD v4) |
| --- | --- | --- | --- | --- |
| Family 1<br>Individual 1 | Homozygous | 1:39322647 | c.4069G>A, p.E1357K | 1.239E-6 (0 hmz) |
| Family 2<br>Individual 2 | Homozygous | 1:39357705 | c.11755C>T, p.R3919W | 7.435E-5 (0 hmz) |
| Family 2<br>Individual 3 | Homozygous | 1:39357705 | c.11755C>T, p.R3919W | 7.435E-5 (0 hmz) |
| Family 3<br>Individual 4 | Homozygous | 1:39444761 | c.19531T>A, p.S6511T | 9.912E-6 (0 hmz) |
| Family 4<br>Individual 5 | Homozygous | 1:39434586 | c.17738C>T, p.P5913L | 2.106E-5 (0 hmz) |
| Family 5<br>Individual 6 | Compound<br>Heterozygous | 1:39422372 | c.15817-2A>G | 1.859E-6 (0 hmz) |
|  |  | 1:39448679 | c.20174T>C, p.L6725P | 6.197E-7 (0 hmz) |
| Family 5<br>Individual 7 | Compound<br>Heterozygous | 1:39422372 | c.15817-2A>G | 1.859E-6 (0 hmz) |
|  |  | 1:39448679 | c.20174T>C, p.L6725P | 6.197E-7 (0 hmz) |
| Family 6<br>Individual 8 | Compound<br>Heterozygous | 1:39257970 | c.470C>T, p.T157I | 1.053E-5 (0 hmz) |
|  |  | 1:39357693 | c.11743T>C, p.S3915P | 1.228E-3 (3 hmz) |
| Family 7<br>Individual 9 | Homozygous | 1:39322623 | c.4045C>A, p.L1349M | 1.107E-3 (2 hmz) |
| Family 8<br>Individual 10 | Heterozygous | 1:39424138-39424140 | c.16261_16262del, p.Q5421Afs*5 | 0 (0 hmz) |

| **CADD PHRED score** | **SpliceAI** | **Primate AI** | **GERP++** | **NMD-EscPredictor** | **ACMG Classification** |
| --- | --- | --- | --- | --- | --- |
| 24.2 | - | Deleterious (0.82) | 5.92 | - | VUS (PM2, PP2) |
| 21.5 | - | Uncertain (0.52) | 6.07 | - | VUS (PM2, PP2) |
| 21.5 | - | Uncertain (0.52) | 6.07 | - | VUS (PM2, PP2) |
| 20.4 | - | Uncertain (0.54) | 6.116 | - | VUS (PM2, PP2) |
| 22 | - | Uncertain (0.74) | 5.8 | - | VUS (PM2, PP2) |
| 35 | 0.99 (AL) | - | - | - | VUS (PM2, PVS1) |
| 23.6 | - | Uncertain (0.68) | 6.17 | - | VUS (PM2, PP2) |
| 35 | 0.99 (AL) | - | - | - | VUS (PM2, PVS1) |
| 23.6 | - | Uncertain (0.68) | 6.17 | - | VUS (PM2, PP2) |
| 23.4 | - | Deleterious (0.9) | 5.41 | - | VUS (PM2, PP3, PP2) |
| 19.12 | - | Benign (0.034) | 6.07 | - | Benign (PP2, BS1, BS2, BP4) |
| 26 | - | Uncertain (0.78) | 5.99 | - | Benign (PP2, BS1, BS2, BP6) |
| - | - | - | - | Degraded by NMD | LP (PVS1,PM2) |

### Phenotypic spectrum of *MACF1* variation

While monoallelic variants affecting the GAR domain in *MACF1* associate well with Lissencephaly 9 with Complex Brainstem Malformations [MIM 618325]^14^, we observe variants (mono– and biallelic) in other domains of *MACF1* that show a partially overlapping yet broader phenotypic spectrum. This makes assessment of domain-specific *MACF1* variation and disease more difficult. To systematically explore the phenotypic spectrum of *MACF1*, deep phenotypic data were collected from collaborating clinicians using a standardized template. Through an inhouse developed tool, RAG-HPO^30^, that uses large language models (LLMs) and Retrieval-Augmented Generation (RAG) followed by manual curation, we comprehensively extracted HPO terms from clinical data and the previously reported cases. Among all 29 affected individuals, we have an average of 16 HPO terms per individual (Figure S1). Gross motor delay, hypotonia, and tethered spinal cord are observed in Individual 8. Since only 3 HPO terms were available from her clinical notes, she was excluded from the quantitative phenotypic analysis, which requires a minimum of 4 HPO terms per case to ensure sufficient phenotypic resolution. Insufficient HPO term annotation can adversely affect clustering performance and the interpretability of metrics such as the gap statistic and Ward evaluation score. Applying the minimum threshold of four HPO terms per case, the analysis yielded a Gap statistic of 0.19 and a Ward evaluation score of 0.865, indicative of moderately structured, well-separated, and internally cohesive clusters.

To contextualize the phenotypic features associated with *MACF1* variants, we included phenotypic data from previously reported disease-causing variants in the GAR domain for comparison with the variants described in this study. Reports of cases with MACF1 variants outside the GAR domain, linked to an expanded phenotype, were also evaluated^11,14,15,17,18,20,21,35^. Furthermore, two cases of unreported inheritance with heterozygous missense *MACF1* variants outside the GAR domain, documented in the DECIPHER database^35^, were incorporated into the analysis. This approach was designed to reduce potential bias from selective case inclusion, providing a balanced and nuanced evaluation of *MACF1*-associated phenotypic variability.

Through HPO-based phenotypic clustering analysis, we can compare and investigate the clinical manifestations of domain-specific variants in *MACF1*. Phenotypic similarity scores generated using the Lin similarity^32^ method are calculated and visualized in a heatmap (Figure 3a). Strikingly, we identify 2 distinct clusters, suggesting that variants outside the GAR domain display traits that have a distinct phenotypic spectrum. Cluster 1 includes the 9 cases described by Dobyns *et al.* with variants in the GAR domain that result in a distinctive phenotype with severe pons hypoplasia and lissencephaly, while cluster 2 includes 19 cases that have a phenotypic spectrum.

**Figure 3:**
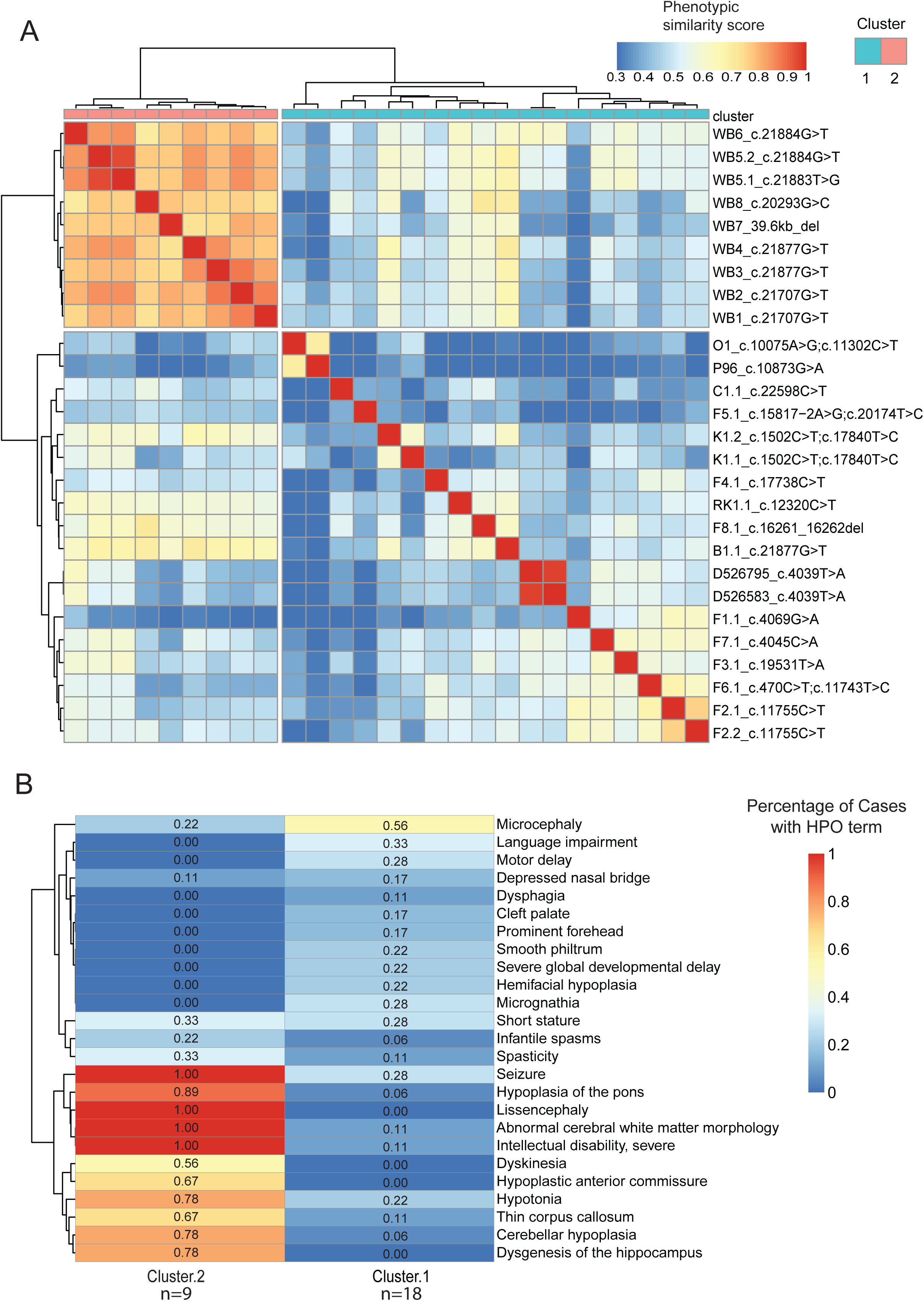
Quantitative phenotypic analyses. (A) Phenotypic similarity heatmap for individuals with *MACF1* variants. A heatmap was generated using proband phenotypic similarity scores and ordered based on Hierarchical Agglomerative Clustering of proband phenotypic similarity. Dendrograms showing clusters are present at the left and top sides of the heatmap. Highest phenotypic similarity score of 1 is shown in red that runs along as the diagonal. Two distinct clusters are noted suggesting clinical subgroups within *MACF1*-related disorder. (B) Significant HPO terms differentiating the 2 clusters is plotted. Applying a generalized linear model (GLM) using a quasipoisson distribution, unique HPO terms significantly associated with the two specific clusters were identified. Percentages of cases in each cluster for each of the significant HPO term is denoted in the columns.

To identify unique HPO terms significantly associated with one of the two specific clusters, we applied a generalized linear model (GLM) using a quasipoisson distribution. This model evaluates the statistical association between HPO terms and cluster assignments, enabling the identification of terms that are most characteristic of each cluster. The heatmap (Figure 3b) highlights distinct phenotypic profiles between the two clusters, showcasing the variability in phenotype prevalence, while considering the number of cases in each cluster. Neurological phenotypes such as lissencephaly, seizures, and severe intellectual disability are notably more prevalent in cases with *MACF1* variants in the GAR domain, while craniofacial and skeletal phenotypes such as microcephaly, cleft palate, micrognathia, and clinodactyly are present in cases with *MACF1* variants outside the GAR domain. Traits like short stature, spasticity, and infantile spasms appear in both clusters, albeit at lower prevalence than traits exhibiting greater variability between clusters, suggesting they may reflect shared but less penetrant features of the overall phenotype. Hierarchical clustering further elucidates patterns of phenotypic co-occurrence, hinting at shared genetic or developmental pathways that may underlie certain phenotype groupings. This approach integrates statistical modeling with quantitative phenotypic clustering to uncover phenotypic patterns and cluster dependencies, providing valuable insights into disease phenotypes and their genetic underpinnings.

### Distinct phenotypic clustering of *MACF1* variants based on domain localization

Phenotypic clustering analysis between these cases and matching OMIM gene-associated HPO terms sets (i.e. ‘gene-level’ analyses, p <0.001, Figure 4a) revealed that variants outside the GAR domain display traits with a distinct phenotypic spectrum. Overall, these analyses provide evidence that variants outside the GAR domain are associated with phenotypes that are more variable, with some overlap of neurologic features associated with variants within the GAR domain. Of note, OMIM phenotypes associated with *NCAPD3* cluster with cases harboring *MACF1* variants outside the GAR domain. Biallelic variants in *NCAPD3* are associated with primary microcephaly-22 (MCPH22), poor overall growth, and variable neurologic features including moderate developmental delay, seizures, and limb hypertonia, illustrating the association of biallelic variation in our *MACF1* cohort with expanded clinical presentations^36^. Although their shared phenotypes might suggest a common pathway or co-expression network, there is currently no direct evidence supporting functional overlap or interaction.

**Figure 4:**
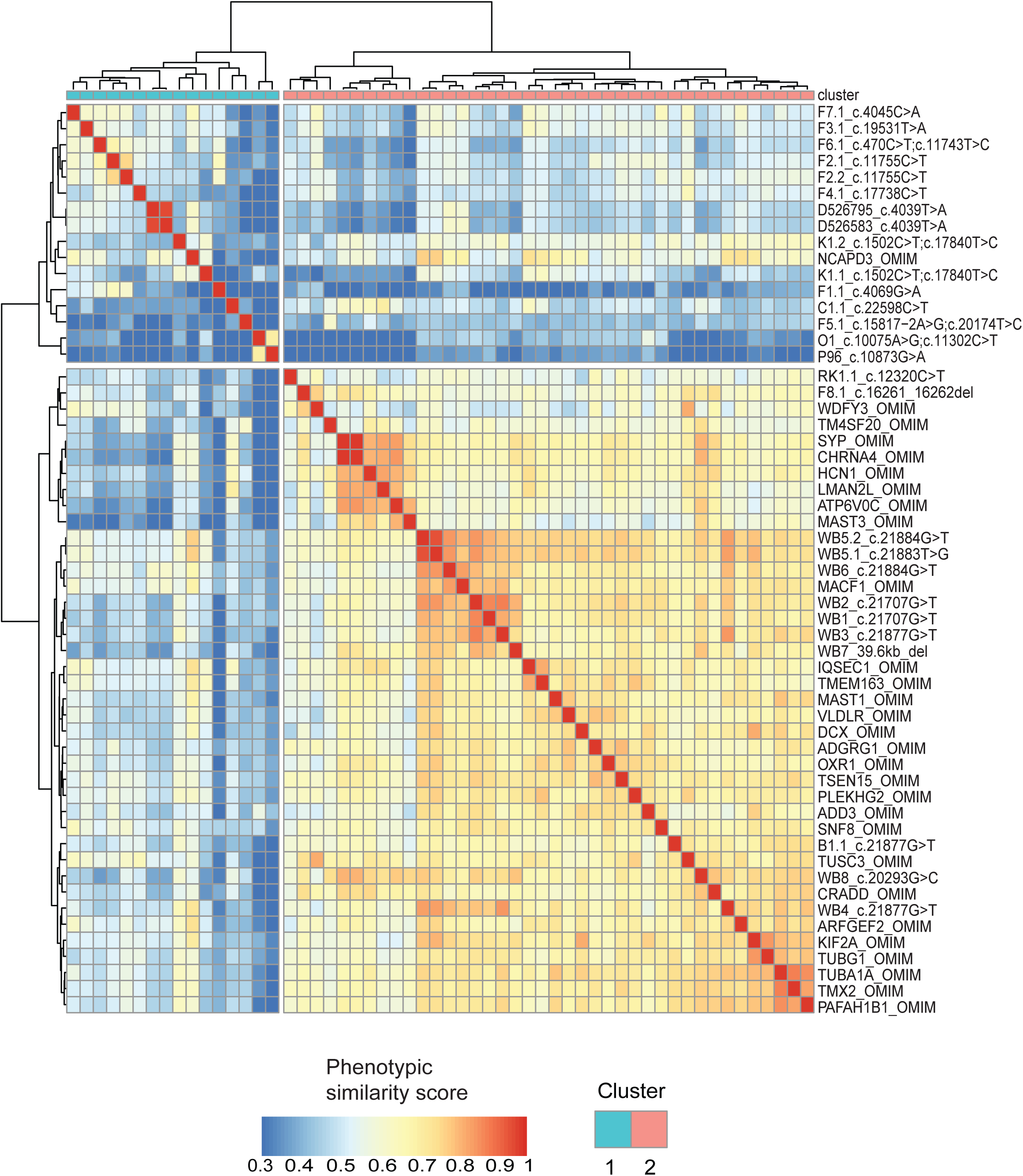
Comparative phenotypic analysis using OMIM gene term sets. Phenotypic similarity heatmap of individuals with *MACF1* variants, analyzed alongside OMIM gene term sets (p < 0.001), reveals two distinct clustering patterns. Cluster 1 comprises cases with MACF1 variants outside the GAR domain, along with *NCAPD3*, which are linked to microcephaly and global developmental delay as shared phenotypes. Cluster 2 includes cases with MACF1 variants within the GAR domain, as well as OMIM genes associated with lissencephaly and cerebellar hypoplasia, consistent with microtubule-binding defects.

The OMIM entry for *MACF1* clusters with the Dobyns *et al.* cases^14^, possibly reflecting the core clinical presentation previously described. Moreover, *MACF1* variants within the GAR domain cluster with genes implicated in cytoskeleton organization (*TUBA1A*, *TUBG1*, *PAFAH1B1*, *DCX*), neuronal development (*HCN1*, *CHRNA4*, *SYP*, *DCX*, *MAST1*, *MAST3*), and intracellular transport (*ARFGEF2*, *SNF8*, *IQSEC1*, *TMX2*). This clustering pattern is particularly noteworthy considering the established role of the MACF1 GAR domain in microtubule binding and its critical function in cellular and neuronal processes.

Comparative analysis of phenotypic data using OMIM disease-associated HPO term sets (i.e. ‘disease-level’ analyses, p < 0.001, Figure 5) demonstrated consistent clustering trends. Cases with variants within the GAR domain were observed to cluster with phenotypes linked to lissencephaly and cortical dysplasia with complex brain malformations. Cluster 2 consists of *de novo* heterozygous cases with frameshift variants or large deletions near the GAR domain, which are strongly associated with intellectual disability, developmental delay, and epilepsy phenotypes. Cluster 3 includes cases described in the literature with *MACF1* variants associated with myasthenic syndrome, clustering with other congenital myasthenic syndrome phenotypes. Cluster 4 includes cases from our cohort with *MACF1* variants outside the GAR domain, which do not align with any known OMIM disease phenotype, potentially indicating that these cases represent broader neurodevelopmental phenotypes. The clustering pattern differences reflect how gene-level annotations capture broader phenotypic variation, while disease-level annotations emphasize canonical diagnoses, highlighting the importance of OMIM annotation context in interpreting phenotype-based clustering results.

**Figure 5:**
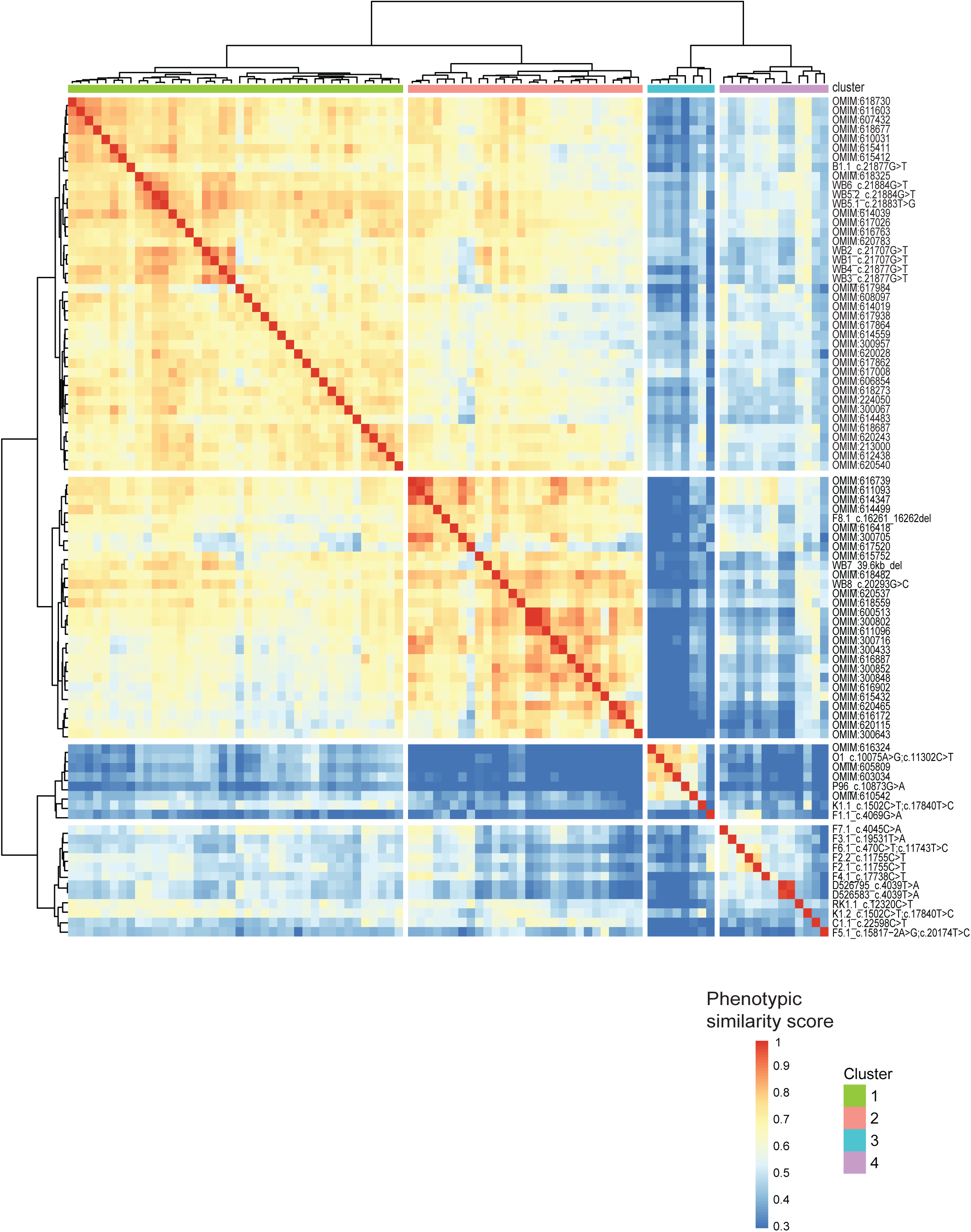
Comparative phenotypic analysis using OMIM disease term sets. Phenotypic similarity heatmap of individuals with *MACF1* variants, analyzed alongside OMIM disease term sets (p < 0.001), reveals four distinct clustering patterns. Cluster 1 includes cases with GAR domain variants that are closely aligned with Lissencephaly and Cortical Dysplasia, Complex, with Other Brain Malformations phenotypes. Cluster 2 includes *de novo* heterozygous cases with frameshift mutations or large deletions near the GAR domain, clustering with intellectual disability, developmental delay, and epilepsy phenotypes. Cluster 3 comprises cases previously described with congenital myasthenic syndrome, which align with myasthenic syndrome phenotypes. Cluster 4 consists of cases from our cohort without a defined OMIM disease phenotype, suggesting that these cases represent broad neurodevelopmental phenotypes.

Overall, *MACF1* variants within the GAR domain demonstrate a strong genotype-phenotype correlation with lissencephaly with brain stem hypoplasia. In contrast, cases with variants outside the GAR domain exhibit a broader neurodevelopmental phenotype with variable expressivity, particularly affecting craniofacial and skeletal features.

## Discussion

This study broadens the understanding of *MACF1*-related disorders by highlighting phenotypic expansion with biallelic or monoallelic variants outside the microtubule-binding GAR domain. Specifically, we describe a constellation of clinical features, including microcephaly, clinodactyly, micrognathia, cleft palate, and skeletal abnormalities, which distinguish these cases from the previously described autosomal dominant (AD) phenotype of Lissencephaly 9 with Complex Brainstem Malformation (MIM#:618325)^14^. Our findings suggest that *MACF1* variants closer to the N-terminal region, encompassing the actin-binding Calponin Homology (CH) domains and cytoskeletal spectrin and plakin repeats, contribute to a more variable autosomal recessive (AR) or autosomal dominant with incomplete penetrance phenotype, implicating distinct structural and functional roles of these domains.

*MACF1* encodes a mega-scaffolding protein critical for cytoskeletal organization, particularly through its ability to cross-link actin and microtubules. This dual interaction is essential for processes such as cell migration, morphology, and division, which are fundamental to neurodevelopment and tissue patterning. Heterozygous missense variants in the GAR domain, which are specifically involved in microtubule binding, have been firmly linked to Lissencephaly 9 with Brainstem Malformation (MIM#:618325), underscoring the importance of microtubule dynamics in neuronal migration^14^. In contrast, the phenotypic features associated with N-terminal *MACF1* variants suggest that perturbations in actin-binding and cytoskeletal anchoring mechanisms contribute to a broader neurodevelopmental impact, along with craniofacial abnormalities and skeletal defects.

The dual phenotypic outcomes linked to *MACF1* variants are reminiscent of domain-specific phenotypes observed in other genes. For instance, pathogenic variants in *COL2A1*, encoding type II collagen, result in distinct disorders^37^. Truncating variants in *COL2A1* cause Stickler syndrome, a connective tissue disorder characterized by ocular, auditory, and skeletal abnormalities, via haploinsufficiency^38,39^. In contrast, spondyloepiphyseal dysplasia, marked by short stature and epiphyseal abnormalities, arises from missense variants in the Gly-X-Y motif, disrupting collagen assembly through a dominant negative effect^38,39^. Similarly, *FGFR1* mutations demonstrate domain-specific effects: variants in the extracellular Ig-like domains cause Pfeiffer syndrome (OMIM: 101600), characterized by craniosynostosis, broad thumbs, and great toes, while mutations in the kinase domain are associated with Kallmann syndrome (OMIM: 147950), distinguished by hypogonadotropic hypogonadism and anosmia^40–42^. These examples highlight how the disruption of specific functional domains of a single gene can give rise to diverse phenotypic outcomes, a concept supported by our findings in *MACF1*.

The presence of biallelic *MACF1* variants in our cohort aligns with established patterns in which dominant and recessive disorders driven by a locus may result in distinct phenotypic spectra and degrees of severity. For instance, *LMNA* variants lead to diverse tissue-specific phenotypes including cardiomyopathy, lipodystrophy, neuropathy, progeria, bone/skin disorders, and syndromes that include combinations of two or more of these phenotypes^43,44^. Like MACF1, LMNA is a large structural protein with critical roles in cellular and nuclear architecture and function. Both *MACF1* and *LMNA* thus share similar complexities in establishing genotype-phenotype correlations such as variability in phenotypic expression from identical variants, the influence of variant location within the gene, the role of post-transcriptional modifications, and the impact of genetic background^45,46^. Such genotype-phenotype correlations emphasize the importance of inheritance patterns and domain-specific functions in shaping clinical outcomes.

Our observations also suggest a potential role for MACF1 in development of the pharyngeal arches and craniofacial structures. While MACF1 has been well-studied in the context of neuronal development, its involvement in craniofacial and skeletal patterning remains unexplored. Given the actin-binding properties of the N-terminal region, it is plausible that disruptions in cytoskeletal anchoring and cell polarity influence pharyngeal pouch development^47^, leading to the craniofacial abnormalities observed in this study. This hypothesis warrants further investigation, particularly in patient cohorts with rare, deleterious non-GAR domain *MACF1* variants.

Complete constitutional loss of *Macf1* in mice results in embryonic lethality prior to somitogenesis, characterized by a failure in primitive streak formation, underscoring the essential developmental role of MACF1^48–50^. In contrast, conditional knockout of *Macf1* in the mouse brain leads to impaired cortical neuron migration, respiratory distress, and early postnatal lethality, highlighting its critical function in neurodevelopment^49^. *MACF1* is classified as highly intolerant to loss-of-function (LoF) variants, with a pLI score of 1 (gnomAD v4) and moderately intolerant to missense variants (z = 6.22)^51,52^. Heterozygous missense variants in the GAR domain are associated with severe phenotypes, including lissencephaly and cortical dysplasia with complex brain malformations, suggesting that these variants may act through an altered function or a dominant-negative mechanism^14^. In contrast, variants outside the GAR domain are more commonly linked to broader neurodevelopmental features, often displaying variable expressivity and incomplete penetrance. Based on these patterns, we hypothesize that non-GAR variants represent hypomorphic alleles resulting in partial LoF and are more likely to follow a recessive mode of inheritance.

A recent study provides valuable insights by identifying the enrichment of *MACF1* variants in epilepsy and their distinct localization^53^. Specifically, their findings suggest that epilepsy is associated with biallelic missense variants in the MACF1 plakin domain, while monoallelic *de novo* variants in the GAR domain associate with lissencephaly 9, and those in spectrin repeats with autism spectrum disorder^53^. Our research builds upon these observations by expanding the recognized phenotypic landscape of *MACF1*-related conditions by integrating detailed clinical characterization, systematic OMIM gene and disease comparisons, and structured HPO-based phenotypic analysis. Notably, we observed variable expressivity and identified individuals with monoallelic variants, who presented with epilepsy, suggesting that *MACF1*-associated phenotypes have complex inheritance patterns. Together, these studies underscore the emerging multifaceted nature of *MACF1*-related neurodevelopmental disorders.

In our cohort, we observe notable intrafamilial variability, with individuals carrying identical biallelic *MACF1* variants exhibiting a range of neurological, skeletal, and craniofacial features. This variability suggests a dosage-sensitive or context-dependent role for MACF1, potentially influenced by genetic or environmental modifiers. It also raises the possibility that a subset of individuals with milder *MACF1*-related neurodevelopmental disorders, particularly older individuals who have not undergone genetic testing, may be underdiagnosed. Further investigation into intrafamilial phenotypic heterogeneity is warranted to refine the clinical spectrum and improve genetic counseling and expectant management for families.

In our phenotypic clustering analysis, we observed notable differences in alignment when comparing phenotypic data from our cases to gene-level (Figure 4) versus disease-level (Figure 5) OMIM annotations. These differences reflect how phenotypic annotations are curated and structured. Gene-based profiles capture broader phenotypic variability, often aggregating phenotypes across multiple associated conditions, whereas disease-level annotations are constrained to specific clinical and molecular diagnoses. This contrast is influenced by varying curation depth and the one-to-many relationship between genes and diseases. As a result, clustering of our MACF1 cases showed stronger alignment with gene-level annotations, especially in the context of phenotypic variability or non-classical clinical features. Additionally, since a single gene can underlie multiple clinically distinct conditions, alignment to gene-level phenotypes may better reflect underlying molecular mechanisms, while disease-level alignment may support specific diagnostic interpretations. Together, these findings highlight the complementary value of gene– and disease-based phenotypic comparisons and underscore the need for careful interpretation of phenotypic clustering results in the context of rare diseases.

In summary, our study expands the phenotypic spectrum of *MACF1*-related disorders and underscores the significance of domain-specific functions in shaping distinct clinical presentations. By linking biallelic non-GAR domain *MACF1* variants to craniofacial, skeletal abnormalities and neurological disorders, we provide novel insights into *MACF1*’s broader developmental roles and highlight the need for comprehensive phenotypic and genetic analyses in elucidating the molecular mechanisms underlying these rare conditions. Further studies are essential to explore the full extent of *MACF1* involvement in cytoskeletal regulation and its impact on human development.

## Data Availability

All data described in this study are provided within the article and supplemental material.

## Acknowledgements

We would like to thank all the families and patients for their participation, as well as referring genetic counselors and physicians.

## Funding

This study was supported in part by the US National Human Genome Research Institute (NHGRI) and National Heart, Lung, and Blood Institute to the Baylor-Hopkins Center for Mendelian Genomics (UM1 HG006542, J.R.L.); NHGRI grant as part of the GREGoR Consortium (U01HG011758 to J.E.P., J.R.L., and R.A.G.); NHGRI grant to Baylor College of Medicine Human Genome Sequencing Center (U54HG003273 to R.A.G.). N.G. is supported by the GREGoR Consortium Research Grant. B.V is supported by the German Research Foundation (DFG) via VO 2138/7-1 grant 469177153, the Heisenberg program VO 2138/8-1 grant 543719215, and the Collaborative Research Center 1690 (A03). P.L.B.C. is supported by the Else Hirschberg womeńs promotion program D.P. is supported by NINDS 1K23 NS125126-01A1and the Doris Duke Charitable Foundation (#2023-0235). Research reported in this publication was supported by the National Institute of Neurological Disorders and Stroke of the National Institutes of Health under Award Numbers U01HG007708 and U01HG007942. The content is solely the responsibility of the authors and does not necessarily represent the official views of the National Institutes of Health.

## Availability of data and materials

All data described in this study are provided within the article and supplemental material.

## Ethics approval and consent to participate

This study adheres to the principles in the Declaration of Helsinki. The study was approved by the Baylor College of Medicine Institutional Review Board, Protocol H-29697. Written informed consent was obtained from all participants as required by the Institutional Review Board. Consent forms are archived and available upon request.

## Author Contributions

Conceptualization: N.G, J.E.P; Data Curation and Analysis: N.G., A.J., J.A.R., D.P., J.E.P.; Resources: J.E.P., J.R.L., R.A.G.; Writing-original draft: N.G.; Writing-review and editing: N.G., A.J., J.A.R., P.B.C., J.A.B., D.B., T.B., I.C., P.DS., J.F., S.G., T.H, I.H., U.U.I, S.N.J, I.J., J.L., E.M., R.M., O.L.R., C.T., T.T., B.V., R.M.Z., I.M.W., K.W., D.P., R.A.G., J.R.L., J.E.P.; Supervision: J.E.P.

## Competing interests

James R. Lupski is a consultant for Genome International. Jennifer E. Posey serves on the Scientific Advisory Board of MaddieBio. Davut Pehlivan provides consulting service for Ionis Pharmaceuticals and Acadia Pharmaceuticals. The Department of Molecular and Human Genetics at Baylor College of Medicine receives revenue from clinical genetic testing completed at Baylor Genetics Laboratory. The remaining authors declare that they have no competing interests.

## Undiagnosed Diseases Network

Alyssa A. Tran, Arjun Tarakad, Ashok Balasubramanyam, Brendan H. Lee, Carlos A. Bacino, Daryl A. Scott, Elaine Seto, Gary D. Clark, Hongzheng Dai, Hsiao-Tuan Chao, Ivan Chinn, James P. Orengo, Jennifer E. Posey, Jill A. Rosenfeld, Kim Worley, Lindsay C. Burrage, Lisa T. Emrick, Lorraine Potocki, Monika Weisz Hubshman, Richard A. Lewis, Ronit Marom, Seema R. Lalani, Shamika Ketkar, Tiphanie P. Vogel, William J. Craigen, Jared Sninsky, Lauren Blieden, Sandesh Nagamani, Hugo J. Bellen, Michael F. Wangler, Oguz Kanca, Shinya Yamamoto, Christine M. Eng, Patricia A. Ward, Pengfei Liu, Adeline Vanderver, Cara Skraban, Edward Behrens, Gonench Kilich, Kathleen Sullivan, Kelly Hassey, Ramakrishnan Rajagopalan, Rebecca Ganetzky, Vishnu Cuddapah, Anna Raper, Daniel J. Rader, Giorgio Sirugo, Vaidehi Jobanputra, Allyn McConkie-Rosell, Kelly Schoch, Mohamad Mikati, Nicole M. Walley, Rebecca C. Spillmann, Vandana Shashi, Alan H. Beggs, Calum A. MacRae, David A. Sweetser, Deepak A. Rao, Edwin K. Silverman, Elizabeth L. Fieg, Frances High, Gerard T. Berry, Ingrid A. Holm, J. Carl Pallais, Joan M. Stoler, Joseph Loscalzo, Lance H. Rodan, Laurel A. Cobban, Lauren C. Briere, Matthew Coggins, Melissa Walker, Richard L. Maas, Susan Korrick, Jessica Douglas, Cecilia Esteves, Emily Glanton, Isaac S. Kohane, Kimberly LeBlanc, Rachel Mahoney, Shamil R. Sunyaev, Shilpa N. Kobren, Brett H. Graham, Erin Conboy, Francesco Vetrini, Kayla M. Treat, Khurram Liaqat, Lili Mantcheva, Stephanie M. Ware, Breanna Mitchell, Brendan C. Lanpher, Devin Oglesbee, Eric Klee, Filippo Pinto e Vairo, Ian R. Lanza, Kahlen Darr, Lindsay Mulvihill, Lisa Schimmenti, Queenie Tan, Surendra Dasari, Abdul Elkadri, Brett Bordini, Donald Basel, James Verbsky, Julie McCarrier, Michael Muriello, Michael Zimmermann, Adriana Rebelo, Carson A. Smith, Deborah Barbouth, Guney Bademci, Joanna M. Gonzalez, Kumarie Latchman, LéShon Peart, Mustafa Tekin, Nicholas Borja, Stephan Zuchner, Stephanie Bivona, Willa Thorson, Herman Taylor, Rakale C. Quarells, Ayuko Iverson, Bruce Gelb, Charlotte Cunningham-Rundles, Eric Gayle, Joanna Jen, Louise Bier, Mafalda Barbosa, Manisha Balwani, Mariya Shadrina, Rachel Evard, Saskia Shuman, Susan Shin, Andrea Gropman, Barbara N. Pusey Swerdzewski, Camilo Toro, Colleen E. Wahl, Donna Novacic, Ellen F. Macnamara, John J. Mulvihill, Maria T. Acosta, Precilla D’Souza, Valerie V. Maduro, Ben Afzali, Ben Solomon, Cynthia J. Tifft, David R. Adams, Elizabeth A. Burke, Francis Rossignol, Heidi Wood, Jiayu Fu, Joie Davis, Leoyklang Petcharet, Lynne A. Wolfe, Margaret Delgado, Marie Morimoto, Marla Sabaii, MayChristine V. Malicdan, Neil Hanchard, Orpa Jean-Marie, Wendy Introne, William A. Gahl, Yan Huang, Andrew Stergachis, Danny Miller, Elisabeth Rosenthal, Elizabeth Blue, Elsa Balton, Emily Shelkowitz, Eric Allenspach, Fuki M. Hisama, Gail P. Jarvik, Ghayda Mirzaa, Ian Glass, Kathleen A. Leppig, Katrina Dipple, Mark Wener, Martha Horike-Pyne, Michael Bamshad, Peter Byers, Runjun Kumar, Seth Perlman, Sirisak Chanprasert, Virginia Sybert, Wendy Raskind, Nitsuh K. Dargie, Chun-Hung Chan, Dr. Francisco Bustos velasq, Isum Ward, Jason Schend, Jennifer Morgan, Megan Bell, Miranda Leitheiser, Mohamad Saifeddine, Paul Berger, Rachel Li, Taylor Beagle, Alexander Miller, Beatriz Anguiano, Beth A. Martin, Brianna Tucker, Chloe M. Reuter, Devon Bonner, Elijah Kravets, Hector Rodrigo Mendez, Holly K. Tabor, Jacinda B. Sampson, Jason Hom, Jennefer N. Kohler, Jennifer Schymick, John E. Gorzynski, Jonathan A. Bernstein, Kevin S. Smith, Laura Keehan, Laurens Wiel, Matthew T. Wheeler, Meghan C. Halley, Mia Levanto, Page C. Goddard, Paul G. Fisher, Rachel A. Ungar, Raquel L. Alvarez, Sara Emami, Shruti Marwaha, Stephen B Montgomery, Suha Bachir, Tanner D Jensen, Taylor Maurer, Terra R. Coakley, Euan A. Ashley, Ali Al-Beshri, Anna Hurst, Brandon M Wilk, Bruce Korf, Elizabeth A Worthey, Kaitlin Callaway, Martin Rodriguez, Tammi Skelton, Tarun KK Mamidi, Andrew B. Crouse, Jordan Whitlock, Mariko Nakano-Okuno, Matthew Might, William E. Byrd, Albert R. La Spada, Changrui Xiao, Elizabeth C. Chao, Eric Vilain, Jose Abdenur, Kirsten Blanco, Maija-Rikka Steenari, Rebekah Barrick, Richard Chang, Sanaz Attaripour, Suzanne Sandmeyer, Tahseen Mozaffar, Alden Huang, Andres Vargas, Bianca E. Russell, Brent L. Fogel, Esteban C. Dell’Angelica, George Carvalho, Julian A. Martínez-Agosto, Layal F. Abi Farraj, Manish J. Butte, Martin G. Martin, Naghmeh Dorrani, Neil H. Parker, Rosario I. Corona, Stanley F. Nelson, Yigit Karasozen, Aaron Quinlan, Alistair Ward, Ashley Andrews, Corrine K. Welt, Dave Viskochil, Erin E. Baldwin, John Carey, Justin Alvey, Laura Pace, Lorenzo Botto, Nicola Longo, Paolo Moretti, Rebecca Overbury, Russell Butterfield, Steven Boyden, Thomas J. Nicholas, Matt Velinder, Gabor Marth, Pinar Bayrak-Toydemir, Rong Mao, Monte Westerfield, Brian Corner, John A. Phillips III, Kimberly Ezell, Lynette Rives, Rizwan Hamid, Serena Neumann, Ashley McMinn, Joy D. Cogan, Thomas Cassini, Alex Paul, Dana Kiley, Daniel Wegner, Erin McRoy, Jennifer Wambach, Kathy Sisco, Patricia Dickson, F. Sessions Cole, Dustin Baldridge, Jimann Shin, Lilianna Solnica-Krezel, Stephen C. Pak, Timothy Schedl, Allen Bale, Carol Oladele, Caroline Hendry, Emily Wang, Hua Xu, Hui Zhang, Lauren Jeffries, María José Ortuño Romero, Mark Gerstein, Michele Spencer-Manzon, Monkol Lek, Nada Derar, Odelya Kaufman, Shrikant Mane, Teodoro Jerves Serrano, Vasilis Vasiliou, Winston Halstead, Yong-Hui Jiang

## Supplementary Figures and Tables

**Figure S1:** Phenotypic depth. The number of HPO terms per individual is plotted. On average, there are 16.7 HPO terms per individual as represented by the black line. The red dotted line marks the minimum threshold of 4 HPO terms required for inclusion in the phenotypic clustering analysis.

**Figure S2:** Gap Statistical Analysis for Determining Optimal Clusters. The gap statistic method compares the variation within observed data clusters against a null reference distribution to identify the optimal number of clusters with meaningful differences. (A) Gap statistic plot for all individuals in our cohort, where the largest gap difference indicates 2 optimal clusters. (B) Gap statistic plot for all individuals combined with OMIM gene term sets (p<0.001), identifying 2 optimal clusters. (C) Gap statistic plot for all individuals combined with OMIM disease term sets (p<0.001), identifying 4 optimal clusters.

**Figure S3:** The absence of heterozygosity (AOH) plot for Individual 1 demonstrates the variant (NM_001394062.1:c.4069G>A, p.(E1357K)) is located within a run of homozygosity (ROH) block of 19.9Mb on chromosome 1, marked by gray zones.

**Table S1:** Detailed summary of MACF1 variants for individuals identified in this study.

**Table S2:** Detailed summary of MACF1 variants for individuals previously reported.

**Table S3:** HPO terms for all individuals in this study.

## Notes

### Author Declarations

The study was approved by the Baylor College of Medicine Institutional Review Board, Protocol H-29697.

## References

1. Gong, T.W., Besirli, C.G., and Lomax, M.I. (2001). *MACF1* gene structure: a hybrid of plectin and dystrophin. Mamm Genome 12, 852–861. 10.1007/s00335-001-3037-3.

2. Hu, L., Su, P., Li, R., Yin, C., Zhang, Y., Shang, P., Yang, T., and Qian, A. (2016). Isoforms, structures, and functions of versatile spectraplakin MACF1. BMB Rep 49, 37–44. 10.5483/BMBRep.2016.49.1.185.

3. Cusseddu, R., Robert, A., and Côté, J.F. (2021). Strength Through Unity: The Power of the Mega-Scaffold MACF1. Front Cell Dev Biol 9, 641727. 10.3389/fcell.2021.641727.

4. Goryunov, D., and Liem, R.K. (2016). Microtubule-Actin Cross-Linking Factor 1: Domains, Interaction Partners, and Tissue-Specific Functions. Methods Enzymol 569, 331–353. 10.1016/bs.mie.2015.05.022.

5. Yang, Y., Dowling, J., Yu, Q.C., Kouklis, P., Cleveland, D.W., and Fuchs, E. (1996). An essential cytoskeletal linker protein connecting actin microfilaments to intermediate filaments. Cell 86, 655–665. 10.1016/s0092-8674(00)80138-5.

6. Leung, C.L., Sun, D., Zheng, M., Knowles, D.R., and Liem, R.K. (1999). Microtubule actin cross-linking factor (MACF): a hybrid of dystonin and dystrophin that can interact with the actin and microtubule cytoskeletons. J Cell Biol 147, 1275–1286. 10.1083/jcb.147.6.1275.

7. Su, P., Tian, Y., Yin, C., Wang, X., Li, D., Yang, C., Pei, J., Deng, X., King, S., Li, Y., and Qian, A. (2022). MACF1 promotes osteoblastic cell migration by regulating MAP1B through the GSK3beta/TCF7 pathway. Bone 154, 116238. 10.1016/j.bone.2021.116238.

8. Ka, M., Jung, E.M., Mueller, U., and Kim, W.Y. (2014). MACF1 regulates the migration of pyramidal neurons via microtubule dynamics and GSK-3 signaling. Dev Biol 395, 4–18. 10.1016/j.ydbio.2014.09.009.

9. Ghasemizadeh, A., Christin, E., Guiraud, A., Couturier, N., Abitbol, M., Risson, V., Girard, E., Jagla, C., Soler, C., Laddada, L., et al. (2021). MACF1 controls skeletal muscle function through the microtubule-dependent localization of extra-synaptic myonuclei and mitochondria biogenesis. Elife 10. 10.7554/eLife.70490.

10. Zhao, F., Ma, X., Qiu, W., Wang, P., Zhang, R., Chen, Z., Su, P., Zhang, Y., Li, D., Ma, J., et al. (2020). Mesenchymal MACF1 Facilitates SMAD7 Nuclear Translocation to Drive Bone Formation. Cells 9. 10.3390/cells9030616.

11. Oury, J., Liu, Y., Töpf, A., Todorovic, S., Hoedt, E., Preethish-Kumar, V., Neubert, T.A., Lin, W., Lochmüller, H., and Burden, S.J. (2019). MACF1 links Rapsyn to microtubule– and actin-binding proteins to maintain neuromuscular synapses. J Cell Biol 218, 1686–1705. 10.1083/jcb.201810023.

12. Chen, H.J., Lin, C.M., Lin, C.S., Perez-Olle, R., Leung, C.L., and Liem, R.K. (2006). The role of microtubule actin cross-linking factor 1 (MACF1) in the Wnt signaling pathway. Genes Dev 20, 1933–1945. 10.1101/gad.1411206.

13. Jørgensen, L.H., Mosbech, M.B., Færgeman, N.J., Graakjaer, J., Jacobsen, S.V., and Schrøder, H.D. (2014). Duplication in the microtubule-actin cross-linking factor 1 gene causes a novel neuromuscular condition. Sci Rep 4, 5180. 10.1038/srep05180.

14. Dobyns, W.B., Aldinger, K.A., Ishak, G.E., Mirzaa, G.M., Timms, A.E., Grout, M.E., Dremmen, M.H.G., Schot, R., Vandervore, L., van Slegtenhorst, M.A., et al. (2018). *MACF1* Mutations Encoding Highly Conserved Zinc-Binding Residues of the GAR Domain Cause Defects in Neuronal Migration and Axon Guidance. The American Journal of Human Genetics 103, 1009–1021. 10.1016/j.ajhg.2018.10.019.

15. Bölsterli, B.K., Steindl, K., Kottke, R., Steinfeld, R., and Boltshauser, E. (2021). Lissencephaly with Brainstem Hypoplasia and Dysplasia: Think MACF1. Neuropediatrics 52, 227. 10.1055/s-0040-1722690.

16. Cox, A.J., Grady, F., Velez, G., Mahajan, V.B., Ferguson, P.J., Kitchen, A., Darbro, B.W., and Bassuk, A.G. (2019). In trans variant calling reveals enrichment for compound heterozygous variants in genes involved in neuronal development and growth. Genetics Research 101, e8, e8. 10.1017/S0016672319000065.

17. Kang, L., Liu, Y., Jin, Y., Li, M., Song, J., Zhang, Y., Zhang, Y., and Yang, Y. (2020). Mutations of MACF1, Encoding Microtubule-Actin Crosslinking-Factor 1, Cause Spectraplakinopathy. Frontiers in Neurology 10. 10.3389/fneur.2019.01335.

18. Polavarapu, K., Sunitha, B., Töpf, A., Preethish-Kumar, V., Thompson, R., Vengalil, S., Nashi, S., Bardhan, M., Sanka, S.B., Huddar, A., et al. (2024). Clinical and genetic characterisation of a large Indian congenital myasthenic syndrome cohort. Brain 147, 281–296. 10.1093/brain/awad315.

19. Bazazzadegan, N., Babanejad, M., Banihashemi, S., Arzhangi, S., Kahrizi, K., Booth, K.T., and Najmabadi, H. (2025). A Novel Candidate Gene *MACF1* is Associated with Autosomal Dominant Non-syndromic Hearing Loss in an Iranian Family. Arch Iran Med 28, 63–66. 10.34172/aim.31746.

20. Capisizu, A., Sandu, C., Caragea, R.M., and Capisizu, A.S. (2024). A missense mutation in the *MACF1* gene in a patient with autism spectrum disorder and epilepsy. J Med Life 17, 1023–1029. 10.25122/jml-2024-0312.

21. Kessler, R., McManus, M., Schmidt, S., Teixeira, S.R., Reynoso Santos, F.J., and Agarwal, S. (2025). A Novel *MACF1* Gene Mutation: Expanding the Fetal and Neonatal Phenotype. Pediatr Neurol 164, 78–80. 10.1016/j.pediatrneurol.2025.01.007.

22. Han, M.-R., Han, K.-M., Kim, A., Kang, W., Kang, Y., Kang, J., Won, E., Tae, W.-S., Cho, Y., and Ham, B.-J. (2019). Whole-exome sequencing identifies variants associated with structural MRI markers in patients with bipolar disorders. Journal of Affective Disorders 249, 159–168. 10.1016/j.jad.2019.02.028.

23. Wang, Q., Li, M., Yang, Z., Hu, X., Wu, H.M., Ni, P., Ren, H., Deng, W., Li, M., Ma, X., et al. (2015). Increased co-expression of genes harboring the damaging de novo mutations in Chinese schizophrenic patients during prenatal development. Sci Rep 5, 18209. 10.1038/srep18209.

24. Kenny, E.M., Cormican, P., Furlong, S., Heron, E., Kenny, G., Fahey, C., Kelleher, E., Ennis, S., Tropea, D., Anney, R., et al. (2014). Excess of rare novel loss-of-function variants in synaptic genes in schizophrenia and autism spectrum disorders. Molecular Psychiatry 19, 872–879. 10.1038/mp.2013.127.

25. Wang, X., Li, N., Xiong, N., You, Q., Li, J., Yu, J., Qing, H., Wang, T., Cordell, H.J., Isacson, O., et al. (2017). Genetic Variants of Microtubule Actin Cross-linking Factor 1 (*MACF1*) Confer Risk for Parkinson’s Disease. Mol Neurobiol 54, 2878–2888. 10.1007/s12035-016-9861-y.

26. Sobreira, N., Schiettecatte, F., Valle, D., and Hamosh, A. (2015). GeneMatcher: a matching tool for connecting investigators with an interest in the same gene. Hum Mutat 36, 928–930. 10.1002/humu.22844.

27. Ramoni, R.B., Mulvihill, J.J., Adams, D.R., Allard, P., Ashley, E.A., Bernstein, J.A., Gahl, W.A., Hamid, R., Loscalzo, J., McCray, A.T., et al. (2017). The Undiagnosed Diseases Network: Accelerating Discovery about Health and Disease. Am J Hum Genet 100, 185–192. 10.1016/j.ajhg.2017.01.006.

28. Gargano, M.A., Matentzoglu, N., Coleman, B., Addo-Lartey, E.B., Anagnostopoulos, A.V., Anderton, J., Avillach, P., Bagley, A.M., Bakštein, E., Balhoff, J.P., et al. (2024). The Human Phenotype Ontology in 2024: phenotypes around the world. Nucleic Acids Res 52, D1333–d1346. 10.1093/nar/gkad1005.

29. Richards, S., Aziz, N., Bale, S., Bick, D., Das, S., Gastier-Foster, J., Grody, W.W., Hegde, M., Lyon, E., Spector, E., et al. (2015). Standards and guidelines for the interpretation of sequence variants: a joint consensus recommendation of the American College of Medical Genetics and Genomics and the Association for Molecular Pathology. Genet Med 17, 405–424. 10.1038/gim.2015.30.

30. Garcia, B.T., Westerfield, L., Yelemali, P., Gogate, N., Andres Rivera-Munoz, E., Du, H., Dawood, M., Jolly, A., Lupski, J.R., and Posey, J.E. (2024). Improving Automated Deep Phenotyping Through Large Language Models Using Retrieval Augmented Generation. medRxiv. 10.1101/2024.12.01.24318253.

31. Greene, D., Richardson, S., and Turro, E. (2017). ontologyX: a suite of R packages for working with ontological data. Bioinformatics 33, 1104–1106.

32. Lin, D. (1998). An information-theoretic definition of similarity. In 1998. pp. 296–304.

33. Rentzsch, P., Witten, D., Cooper, G.M., Shendure, J., and Kircher, M. (2019). CADD: predicting the deleteriousness of variants throughout the human genome. Nucleic Acids Res 47, D886–d894. 10.1093/nar/gky1016.

34. Davydov, E.V., Goode, D.L., Sirota, M., Cooper, G.M., Sidow, A., and Batzoglou, S. (2010). Identifying a High Fraction of the Human Genome to be under Selective Constraint Using GERP++. PLOS Computational Biology 6, e1001025. 10.1371/journal.pcbi.1001025.

35. Firth, H.V., Richards, S.M., Bevan, A.P., Clayton, S., Corpas, M., Rajan, D., Vooren, S.V., Moreau, Y., Pettett, R.M., and Carter, N.P. (2009). DECIPHER: Database of Chromosomal Imbalance and Phenotype in Humans Using Ensembl Resources. The American Journal of Human Genetics 84, 524–533. 10.1016/j.ajhg.2009.03.010.

36. Martin, C.A., Murray, J.E., Carroll, P., Leitch, A., Mackenzie, K.J., Halachev, M., Fetit, A.E., Keith, C., Bicknell, L.S., Fluteau, A., et al. (2016). Mutations in genes encoding condensin complex proteins cause microcephaly through decatenation failure at mitosis. Genes Dev 30, 2158–2172. 10.1101/gad.286351.116.

37. Nishimura, G., Haga, N., Kitoh, H., Tanaka, Y., Sonoda, T., Kitamura, M., Shirahama, S., Itoh, T., Nakashima, E., Ohashi, H., and Ikegawa, S. (2005). The phenotypic spectrum of *COL2A1* mutations. Hum Mutat 26, 36–43. 10.1002/humu.20179.

38. Deng, H., Huang, X., and Yuan, L. (2016). Molecular genetics of the *COL2A1*-related disorders. Mutation Research/Reviews in Mutation Research 768, 1–13. 10.1016/j.mrrev.2016.02.003.

39. Barat-Houari, M., Dumont, B., Fabre, A., Them, F.T., Alembik, Y., Alessandri, J.L., Amiel, J., Audebert, S., Baumann-Morel, C., Blanchet, P., et al. (2016). The expanding spectrum of *COL2A1* gene variants IN 136 patients with a skeletal dysplasia phenotype. Eur J Hum Genet 24, 992–1000. 10.1038/ejhg.2015.250.

40. Lansdon, L.A., Bernabe, H.V., Nidey, N., Standley, J., Schnieders, M.J., and Murray, J.C. (2017). The Use of Variant Maps to Explore Domain-Specific Mutations of *FGFR1*. Journal of Dental Research 96, 1339–1345 10.1177/0022034517726496.

41. Dodé, C., Levilliers, J., Dupont, J.M., De Paepe, A., Le Dû, N., Soussi-Yanicostas, N., Coimbra, R.S., Delmaghani, S., Compain-Nouaille, S., Baverel, F., et al. (2003). Loss-of-function mutations in *FGFR1* cause autosomal dominant Kallmann syndrome. Nat Genet 33, 463–465 10.1038/ng1122.

42. Passos-Bueno, M.R., Wilcox, W.R., Jabs, E.W., Sertié, A.L., Alonso, L.G., and Kitoh, H. (1999). Clinical spectrum of fibroblast growth factor receptor mutations. Hum Mutat 14, 115–125. 10.1002/(sici)1098-1004(1999)14:2<115::Aid-humu3>3 0.Co;2-2.

43. Lin, E.W., Brady, G.F., Kwan, R., Nesvizhskii, A.I., and Omary, M.B. (2020). Genotype-phenotype analysis of LMNA-related diseases predicts phenotype-selective alterations in lamin phosphorylation. Faseb j 34, 9051–9073. 10.1096/fj.202000500R.

44. Carboni, N., Politano, L., Floris, M., Mateddu, A., Solla, E., Olla, S., Maggi, L., Antonietta Maioli, M., Piras, R., Cocco, E., et al. (2013). Overlapping syndromes in laminopathies: a meta-analysis of the reported literature. Acta Myol 32, 7–17.

45. Scharner, J., Gnocchi, V.F., Ellis, J.A., and Zammit, P.S. (2010). Genotype-phenotype correlations in laminopathies: how does fate translate? Biochem Soc Trans 38, 257–262 10.1042/bst0380257.

46. Zhao, J., Zhang, H., Pan, C., He, Q., Zheng, K., and Tang, Y. (2024). Advances in research on the relationship between the *LMNA* gene and human diseases (Review). Mol Med Rep 30, 236. 10.3892/mmr.2024.13358.

47. Quinlan, R., Martin, P., and Graham, A. (2004). The role of actin cables in directing the morphogenesis of the pharyngeal pouches. Development 131, 593–599. 10.1242/dev.00950.

48. Wu, X., Kodama, A., and Fuchs, E. (2008). ACF7 Regulates Cytoskeletal-Focal Adhesion Dynamics and Migration and Has ATPase Activity. Cell 135, 137–148. 10.1016/j.cell.2008.07.045.

49. Goryunov, D., He, C.Z., Lin, C.S., Leung, C.L., and Liem, R.K. (2010). Nervous-tissue-specific elimination of microtubule-actin crosslinking factor 1a results in multiple developmental defects in the mouse brain. Mol Cell Neurosci 44, 1–14. 10.1016/j.mcn.2010.01.010.

50. Bernier, G., Mathieu, M., De Repentigny, Y., Vidal, S.M., and Kothary, R. (1996). Cloning and characterization of mouse ACF7, a novel member of the dystonin subfamily of actin binding proteins. Genomics 38, 19–29. 10.1006/geno.1996.0587.

51. Chen, S., Francioli, L.C., Goodrich, J.K., Collins, R.L., Kanai, M., Wang, Q., Alföldi, J., Watts, N.A., Vittal, C., Gauthier, L.D., et al. (2024). A genomic mutational constraint map using variation in 76,156 human genomes. Nature 625, 92–100 10.1038/s41586-023-06045-0.

52. Gudmundsson, S., Singer-Berk, M., Watts, N.A., Phu, W., Goodrich, J.K., Solomonson, M., Rehm, H.L., MacArthur, D.G., and O’Donnell-Luria, A. (2022). Variant interpretation using population databases: Lessons from gnomAD. Hum Mutat 43, 1012–1030. 10.1002/humu.24309.

53. Lei, X.-Y., Zhang, M.-W., Sun, H., Song, W., Liang, X.-Y., Wang, C.-S., Luo, S., Li, B.-M., Liu, X.-R., Wang, Y., et al. (2025). Identification of *MACF1* as a causative gene of generalised epilepsy. Journal of Medical Genetics, jmg-2025–110699. 10.1136/jmg-2025-110699.

