## Supplemental Text for "Domain specific phenotypic expansion associated with variants in *MACF1*"

### Supplementary Appendix

### Supplemental Methods

<u>Exome/Genome sequencing and analysis</u>

Exome sequencing (ES) for Family 1 was performed at the BCM Human Genome Sequencing Center using the VCRome2.1 + PKv2 capture design, with 100 bp paired-end reads generated on the HiSeq2000 platform and analyzed using CASAVA 1.83-v3.6 for alignment and variant calling. Clinical ES (cES) for Family 2 was conducted at both Baylor Genetics (proband) and GeneDx (affected sibling). Family 3 underwent proband-parent trio genome sequencing (GS) at Rady Children's Institute for Genomic Medicine. Through the UDN program, Family 4 underwent ES at Baylor Genetics, while Family 6 had GS performed on fibroblast samples at Baylor Genetics and on blood samples at HudsonAlpha. Families 5 underwent cES at GeneDx. ES for Family 7 was performed at the Institute of Human Genetics, University of Wüzburg and for Family 9 at Timone's Children Hospital APHM, Marseille, France.

### Supplemental Results

<u>Case descriptions</u>

The proband (Individual 1, Family 1), exhibited significant craniofacial and digital dysmorphism, including facial asymmetry, hyperostosis, left central facial paralysis, small crumpled and cupped ears, microstomia, and 5th finger clinodactyly. Additional notable features comprised hirsutism and synophrys (Figure 1, Table 1). Neurological assessments indicated right frontotemporal cortical dysplasia and falx cerebri calcification on brain MRI, alongside focal epileptic discharges in the left frontocentral region on sleep-wake video EEG. Whole-exome sequencing of this individual prioritized a homozygous *MACF1* variant [NM_001394062.1:c.4069G>A, p.(E1357K)] localized within a 19.9 Mb region of Absence of Heterozygosity (Total AOH: 146.8 Mb) (Figure S3).

Individual 2 presented with spastic paraplegia, micrognathia, microcephaly, and profound delays in speech, motor development, and cognition. Brain MRI findings included a thin corpus callosum and reduced cerebral white matter volume. His older brother, Individual 3, exhibited spastic diplegia, attention-deficit/hyperactivity disorder (ADHD), right-sided microtia, and right facial nerve abnormalities leading to facial paralysis. Brain MRI of Individual 3 showed mild periventricular foci of increased FLAIR and T2 signal. Both siblings were found to be homozygous for the *MACF1* variant [NM_001394062.1:c.11755C>T; p.(R3919W)]. Notably, Individual 2 displayed a more severe phenotype compared to Individual 3, despite sharing the identical homozygous variant.

Individual 4 was born full term following a pregnancy notable for decreased fetal movement. At birth, she was small for gestational age, had a cleft palate, and paucity of movement. Developmental delay including motor and speech was noted, as well as growth delay, and severe intellectual disability. At early teenage years, she demonstrated substantial postnatal growth deficiency (weight Z = -4.11, height -2.85, OFC -4.33). Her exam at that time was notable for a repaired cleft palate, bifid uvula, simple cupped ears, bilateral radial head anomalies, distal hypoplasia of fingers with thenar hypoplasia, bulbous toes, and short 5th metatarsals. Her behavior at that time was described as quite hyperactive with frequent elopement. She then began to demonstrate developmental regression characterized by reduced interactiveness, reduced ambulation, and loss of speech. In the setting of a hospitalization for respiratory distress, she required long-term tracheostomy and gastrostomy tube placement. Most recently, she demonstrates limited eye tracking with no spontaneous movement, intermittent low temperature, and hypotension. Evaluations have included a brain MRI that demonstrated global white matter, deep grey nuclei, and cerebellar volume loss. A repeat brain MRI demonstrated further progression of atrophy involving the cerebellum and pons. MRI spectroscopy demonstrated abnormal signals, but without a specific, recognizable pattern. She was found to have a homozygous missense variant in *MACF1*, [NM_001394062.1:c.19531T>A, p.S6511T] predicted to be damaging (CADD PHRED 20.4[1]) and evolutionarily conserved (GERP++ 6.116[2]). Genetic testing of muscle further demonstrated a mitochondrial deletion (m.3259-16069) at 40% heteroplasmy that was not detected in buccal or blood samples. Muscle electron transport chain studies were normal. This mtDNA deletion is suspected to have contributed to her developmental regression, which is not a feature observed in other individuals with non-GAR domain *MACF1* variants. This case represents one of the oldest individuals with non-GAR domain *MACF1* variants and may provide prognostic insight into the long-term outcomes of individuals with similar variants. A recent study reports that MACF1 is expressed throughout the lifespan, with three major peaks in the fetal, early childhood and adulthood stages[3]. These findings further emphasize the need for additional longitudinal data to elucidate MACF1's role in progressive neurological disorders.

Individual 5 has a history of short stature, poor weight gain, borderline microcephaly, dysmorphic features, and developmental delay. There is a family history of consanguinity. Exome sequencing performed through the UDN study identified a homozygous missense variant in *MACF1*, [NM_001394062.1:c.17738C>T (p.P5913L)], located outside the GAR domain. This variant is predicted to be damaging (CADD PHRED 22[1]) and lies in a highly conserved region (GERP++ 5.8[2]). Two additional variants potentially contributing to her phenotype were also identified, including a homozygous in-frame deletion in *POC1A* (c.79_87delTTCAGTATC, p.F27_I29del), classified as a VUS, and an apparently de novo likely pathogenic variant in SCN4A (c.3403C>A, p.R1135S). Biallelic variants in *POC1A* are known to be associated with Short stature, Onychodysplasia, Facial dysmorphism, and Hypotrichosis (SOFT syndrome)[4], while heterozygous missense variants in *SCN4A* are linked to a range of neuromuscular phenotypes, such as congenital myopathy and periodic paralysis[5,6]. Despite the presence of these additional candidate variants, the inclusion of Individual 5 supports a comprehensive investigation into the contribution of non-GAR domain *MACF1* variants to neurodevelopmental phenotypes.

Individual 6 presented with autism and paroxysmal abnormal movements. His brain MRI revealed non-specific T2/FLAIR hyperintensity. His younger sister, Individual 7, exhibited gross motor delay, hypotonia, constipation, and a tethered spinal cord. Her brain MRI was reported as normal. Both siblings were found to be compound heterozygous for *MACF1* variants [NM_001394062.1:c.9631-2A>G; NM_001394062.1:c.13997T>C; p.(L6725P)], with both variants located outside the GAR domain. Parental genetic analysis confirmed the maternal inheritance of the heterozygous c.9631-2A>G *MACF1* variant and paternal inheritance of the heterozygous c.13997T>C *MACF1* variant. A third sibling with autism, was found to carry only the heterozygous c.9631-2A>G *MACF1* variant.

Individual 8 harbors compound heterozygous variants in the non-GAR domain of *MACF1*. The first variant, [NM_001394062.1:c.470C>T (p.T157I)], is inherited from the unaffected mother, demonstrates several features suggestive of pathogenicity, including high evolutionary conservation (GERP++ score 5.41[2]), damaging *in silico* predictions (CADD PHRED 23.4[1]), and rare in population databases (gnomAD v4 AF 1.053E-5[7]); however, this variant is currently classified as a Variant of Uncertain Significance (VUS) according to ACMG/AMP guidelines[8]. The second variant, [NM_001394062.1:c.11743T>C (p.S3915P)], is inherited from the unaffected father, was initially considered ultra-rare (AF 7.96E-4) with no homozygous individuals reported in gnomAD v3. It is classified as likely benign based on increased population frequency (AF 1.228E-3[7]) in gnomAD v4 and the presence of three homozygous individuals. The variant is located in a highly conserved region (GERP++ 6.07[2]). Given this context, we considered a potential modifier effect and both variants were included in our analysis.

Individual 9 carries a homozygous *MACF1* variant, [NM_001394062.1:c.4045C>A (p.L1349M)], currently classified as benign due to the presence of two homozygous individuals in gnomAD v4. At the time of the initial analysis, the variant was considered rare, with no homozygous individuals reported in gnomAD v3 (AF 1.11E-3). Despite its reclassification, the variant resides in a conserved region (GERP++ score 5.99[2]), has a high CADD score (CADD PHRED 26[1]), and is associated with a phenotype consistent with other affected individuals in the cohort, supporting its inclusion in the study.

Individual 10 exhibited a complex clinical presentation including microcephaly, developmental delay, seizures, and significant delays in both speech and motor development. Diagnostic imaging and electrophysiological studies revealed left cortical frontal dysplasia on brain MRI and moderate epileptiform activity in the left frontotemporal region on EEG. Phenotypic assessment also noted coarse facial features and Type 1 diabetes. Genetic analysis via trio exome sequencing identified a de novo heterozygous truncating variant in *MACF1* [NM_001394062.1:c.16261_16262del p.(Gln5421fs)], which is predicted to undergo nonsense-mediated decay, and was included due to the strong phenotypic overlap and the predicted impact of the variant.

References

1. Rentzsch, P., Witten, D., Cooper, G.M., Shendure, J., and Kircher, M. (2019). CADD: predicting the deleteriousness of variants throughout the human genome. Nucleic Acids Res *47*, D886-d894. 10.1093/nar/gky1016.
2. Davydov, E.V., Goode, D.L., Sirota, M., Cooper, G.M., Sidow, A., and Batzoglou, S. (2010). Identifying a High Fraction of the Human Genome to be under Selective Constraint Using GERP++. PLOS Computational Biology *6*, e1001025. 10.1371/journal.pcbi.1001025.
3. Lei, X.-Y., Zhang, M.-W., Sun, H., Song, W., Liang, X.-Y., Wang, C.-S., Luo, S., Li, B.-M., Liu, X.-R., Wang, Y., et al. (2025). Identification of *MACF1* as a causative gene of generalised epilepsy. Journal of Medical Genetics, jmg-2025-110699. 10.1136/jmg-2025-110699.
4. Sarig, O., Nahum, S., Rapaport, D., Ishida-Yamamoto, A., Fuchs-Telem, D., Qiaoli, L., Cohen-Katsenelson, K., Spiegel, R., Nousbeck, J., Israeli, S., et al. (2012). Short Stature, Onychodysplasia, Facial Dysmorphism, and Hypotrichosis Syndrome Is

Caused by a *POC1A* Mutation. The American Journal of Human Genetics *91*, 337-342. 10.1016/j.ajhg.2012.06.003.

5. Brancati, F., Valente, E.M., Davies, N.P., Sarkozy, A., Sweeney, M.G., LoMonaco, M., Pizzuti, A., Hanna, M.G., and Dallapiccola, B. (2003). Severe infantile hyperkalaemic periodic paralysis and paramyotonia congenita: broadening the clinical spectrum associated with the T704M mutation in *SCN4A*. J Neurol Neurosurg Psychiatry *74*, 1339-1341. 10.1136/jnnp.74.9.1339.
6. Cannon, S.C. (2015). Channelopathies of skeletal muscle excitability. Compr Physiol *5*, 761-790. 10.1002/cphy.c140062.
7. Chen, S., Francioli, L.C., Goodrich, J.K., Collins, R.L., Kanai, M., Wang, Q., Alföldi, J., Watts, N.A., Vittal, C., Gauthier, L.D., et al. (2024). A genomic mutational constraint map using variation in 76,156 human genomes. Nature *625*, 92-100. 10.1038/s41586-023-06045-0.
8. Richards, S., Aziz, N., Bale, S., Bick, D., Das, S., Gastier-Foster, J., Grody, W.W., Hegde, M., Lyon, E., Spector, E., et al. (2015). Standards and guidelines for the interpretation of sequence variants: a joint consensus recommendation of the American College of Medical Genetics and Genomics and the Association for Molecular Pathology. Genet Med *17*, 405-424. 10.1038/gim.2015.30.
