## Supplementary figures and images for "Domain specific phenotypic expansion associated with variants in *MACF1*"

### Figure S1

#### Number of HPO Terms per Individual

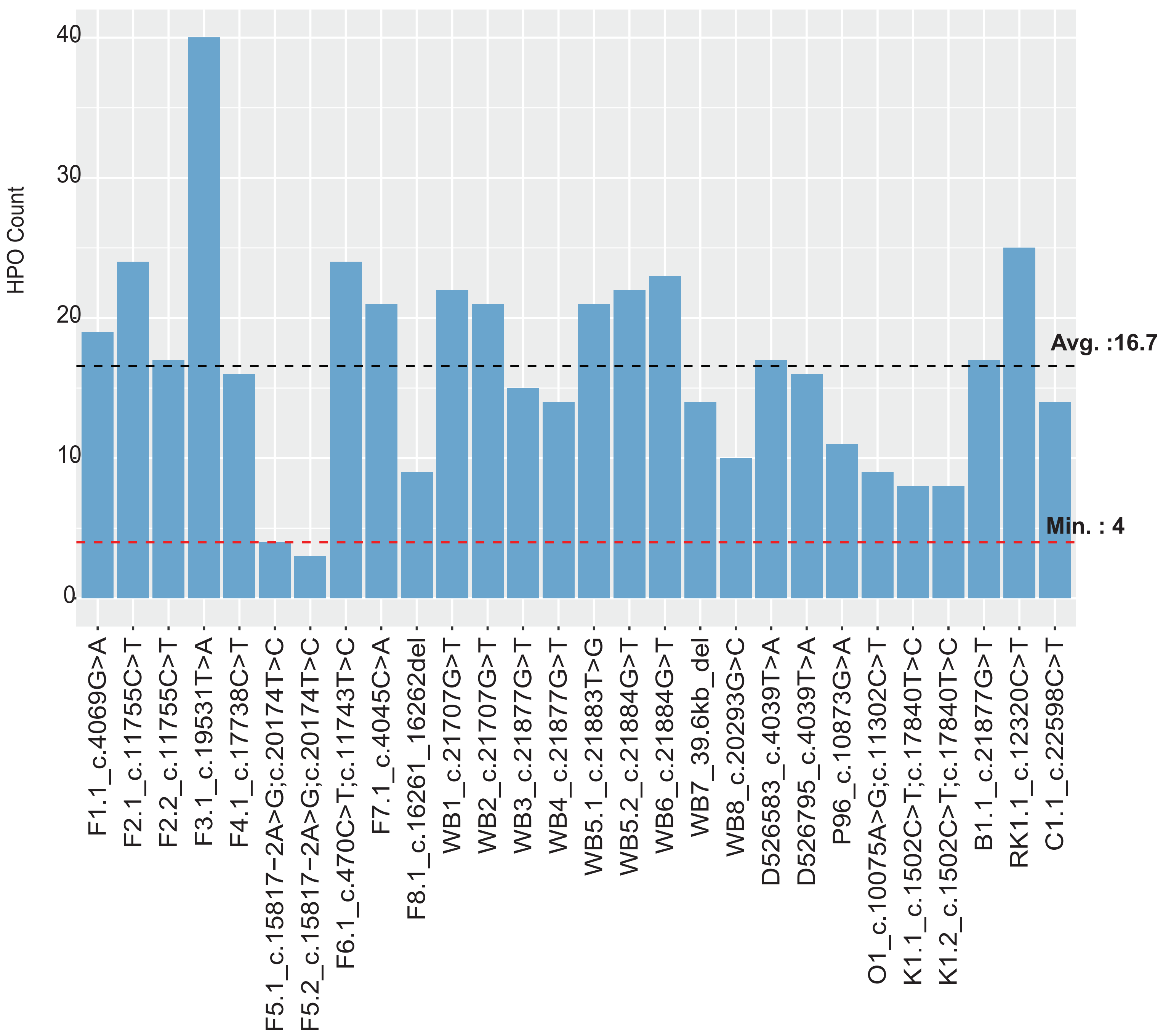

### Figure S2

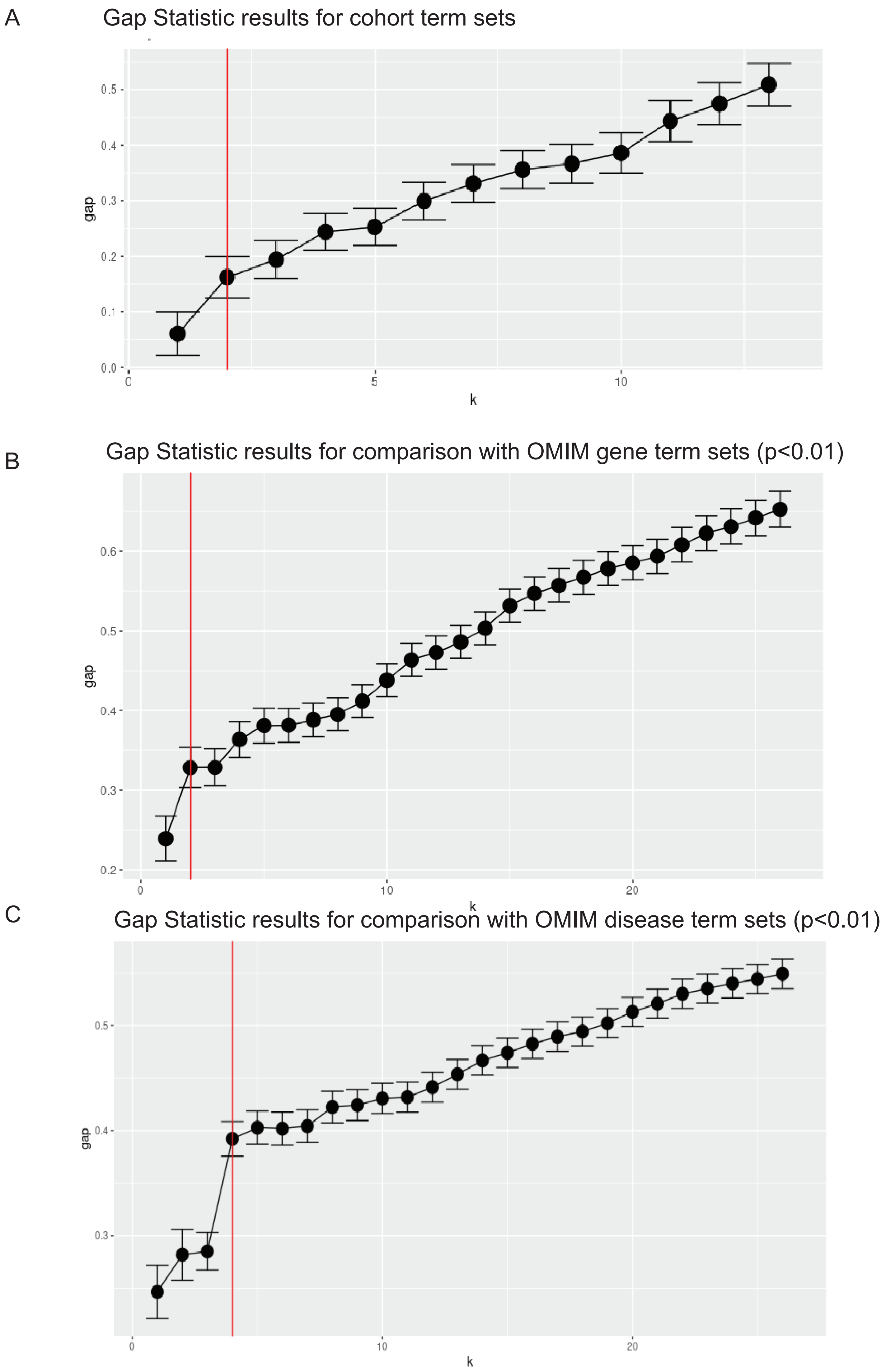

### Figure S3

**Individual 1**
Chr1:39322647
NM_001394062.1:c.4069G>A, p.(E1357K)

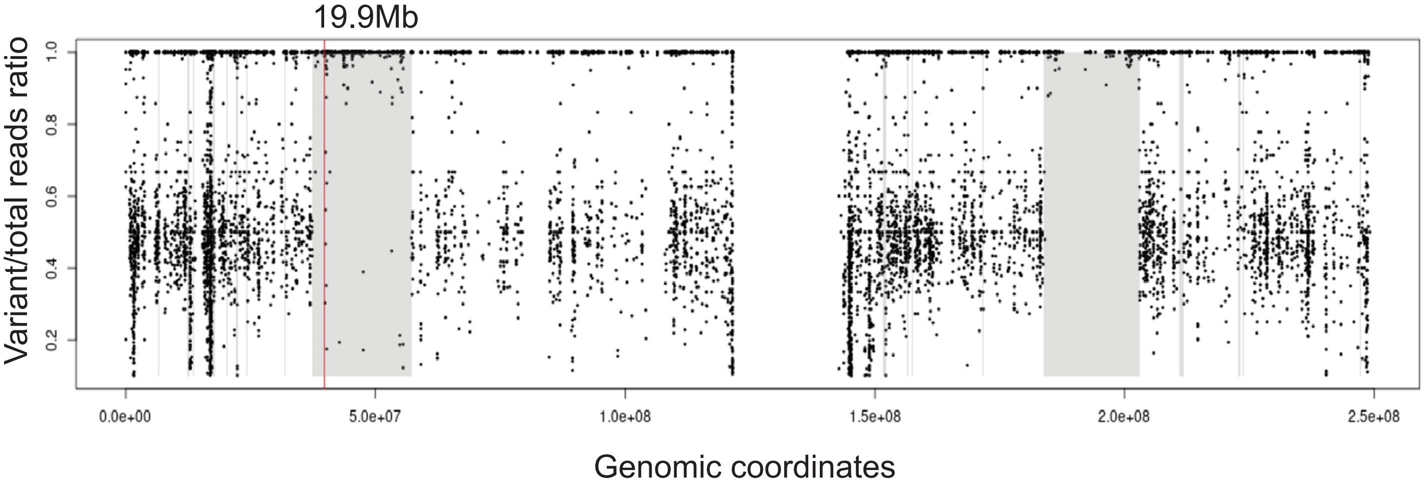
